# Inflammatory reactivity is unrelated to childhood adversity or provoked modulation of nociception

**DOI:** 10.1101/2024.12.16.24319079

**Authors:** Gillian J Bedwell, Luyanduthando Mqadi, Peter Kamerman, Mark R Hutchinson, Romy Parker, Victoria J Madden

**Author notes:** **Corresponding author:** Postal address: D23 Department of Anaesthesia, Groote Schuur Hospital, Anzio Rd, Observatory, 7925, South Africa. **Previous presentation of this work:** Preliminary results were presented in a poster at the 2024 World Congress on Pain, in Amsterdam; 2023 Australasian Neuroscience Society Annual Conference, in Brisbane; and 2023 PainSA Congress, in Johannesburg.

## Abstract

Adversity in childhood elevates the risk of persistent pain in adulthood. Neuroimmune interactions are a candidate mechanistic link between childhood adversity and persistent pain. We aimed to clarify whether immune reactivity is associated with provoked differences in nociceptive processing in adults with a range of childhood adversity. Pain-free adults (n=96; 61 female; median (range) age: 23 (18-65) years old) with a history of mild to severe childhood adversity underwent psychophysical assessments before and after *in vivo* neural provocation (high-frequency electrical stimulation) and, separately, before and after *in vivo* immune provocation (influenza vaccine administration). Psychophysical assessments included the surface area of secondary hyperalgesia after neural provocation and change in conditioned pain modulation (test stimulus: pressure pain threshold; conditioning stimulus: cold water immersion) after immune provocation. Immune reactivity was operationalised as IL-6 and TNF-α expression after *in vitro* lipopolysaccharide provocation of whole blood. We hypothesised associations between immune reactivity and (1) childhood adversity, (2) induced secondary hyperalgesia, and (3) vaccine-associated change in conditioned pain modulation. We found that provoked expression of pro-inflammatory cytokines was not statistically associated with childhood adversity, induced secondary hyperalgesia, or vaccine-associated change in conditioned pain modulation. The current findings from a heterogenous sample cast doubt on two prominent ideas: that childhood adversity primes the inflammatory system for hyper-responsiveness in adulthood and that nociceptive reactivity is linked to inflammatory reactivity. This calls for the broader inclusion of heterogeneous samples in fundamental research to investigate the psychoneuroimmunological mechanisms underlying vulnerability to persistent pain.

## Introduction

A large body of evidence shows that childhood adversity elevates the risk of persistent pain in adulthood [46]. Strikingly, this body of evidence includes no studies from low– and middle-income countries (LMICs) or the African continent, where childhood adversity is disproportionately high [2], adversity is interpreted with diverse cultural perspectives, and social support may enhance resilience [48]. Additionally, genetic diversity [45] and different environmental immune exposures [8] may shape the mechanistic pathways that link childhood adversity to persistent pain in people living in LMICs. Clarifying how childhood adversity increases risk of persistent pain in LMICs is necessary to inform targeted interventions.

Neuroimmune reactivity is a candidate mechanistic link. Adults with a history of childhood adversity display elevated interleukin(IL)-6 and tumour necrosis factor(TNF)-α [14, 20, 60]—a pro-inflammatory profile that may confer vulnerability to persistent pain [34]. Indeed, pro-inflammatory cytokine expression is increased in painful inflammatory conditions such as inflammatory bowel disease [31], rheumatoid arthritis [32], and interstitial cystitis/bladder pain syndrome [58], and predicts the number of painful sites in bladder pain syndrome [58]. Adults with a history of childhood adversity also display heightened amygdala responsiveness and vigilance to threatening stimuli [11, 42, 64, 66], suggesting reduced inhibitory control [47]. Childhood adversity coupled with low socioeconomic status is associated with diminished conditioned pain modulation [63]. Childhood adversity is also associated with greater peaks and slower decay of temporal summation [75]. Reduced conditioned pain modulation and increased temporal summation predict worse pain outcomes at follow-up [17]. Together, these data link childhood adversity to pro-nociceptive modulation that likely elevates vulnerability to persistent pain.

Experimental immune provocations offer an opportunity to capture the functional ‘reactivity’ of the immune system to a standardised stimulus. One useful *in vivo* immune provocation is the influenza vaccine [53]. A matched, *in vitro* immune provocation is achieved by stimulating whole blood with lipopolysaccharide. Whereas the *in vivo* model captures the considerable complexity of immune and cross-system interactions that occur within the dynamic living person, the *in vitro* model may enhance clarity by locking the snapshot of responsiveness to the time of the blood draw, thus allowing tighter inter-individual comparison than a real-life provocation.

Experimental neural provocations offer a comparable opportunity to study the ‘reactivity’ of the neural system to provocation. High-frequency electrical stimulation [21, 29, 30, 51, 65] mimics the nociceptive barrage to the central nervous system after tissue damage without causing actual tissue damage [51]. The resulting, time-limited secondary hyperalgesia is mediated by heterotopic long-term potentiation-like processes in the spinal dorsal horn [30], and can be quantified by the anatomical spread (i.e. surface area) and magnitude of hyperalgesia.

This study aimed to test whether neuroimmune reactivity presents a mechanistic link between childhood adversity and an adult’s vulnerability to persistent pain. This study tested three hypotheses in adults: (1) childhood adversity will predict expression of IL-6 and TNF-α after *in vitro* immune provocation, and (2) provoked expression of IL-6 and TNF-α will predict psychophysical pain-related outcomes after *in vivo* neural provocation, and, separately, (3) after *in vivo* immune provocation.

## Methods

### Study overview

This was a basic experimental study involving humans. The study protocol was approved by the University of Cape Town, Faculty of Health Sciences Human Research Ethics Committee (560/2021), registered at clinicaltrials.gov (NCT06127693), and locked online at Open Science Framework *[insert link at publication]* [37], and we followed the CONSORT reporting guidelines [59] (Supplementary file: Section 1, Table S1). All protocol deviations are explained in Supplementary file: Section 2, Table S2.

Otherwise healthy pain-free adult volunteers, together covering a range of self-reported childhood adversity ratings, underwent a two-visit procedure, starting at similar times on two consecutive mornings. On morning 1, participants had their blood drawn, answered questionnaires, and underwent baseline psychophysical testing. Thereafter, participants were exposed to the neural provocation, and psychophysical testing was repeated. Next, participants received the immune provocation. On morning 2, participants had their blood drawn (results not presented in this report), answered questionnaires, underwent psychophysical testing and exited the study. All participants underwent both neural and immune provocations so that the reactivity of both systems was characterised within each individual. Data were collected from June 2022 to September 2022, and from May 2023 to September 2023 at the University of Cape Town.

### Participants

We recruited pain-free adult volunteers (18 – 65 years old) using posters, social media, and word of mouth. Volunteers received study details via email and were screened for eligibility (Table 1) through an online questionnaire using the REDCap electronic data capture tools hosted at the University of Cape Town [18, 19]. Participants provided written informed consent. Participants could withdraw at any stage during or up to 1 hour after each testing session, with options to retain or destroy their data. Participants were compensated 300 ZAR (∼16 USD) in cash upon procedure completion.

**Table 1:** Exclusion criteria. Reasons for each criterion are specified using crosses in the applicable column. HFS: high-frequency electrical stimulation; IL: interleukin; NSAIDs: non-steroidal anti-inflammatory drugs; TNF: tumour necrosis factor.

| Exclusion criteria | Safety risk |  |  | Confounding risk |  |
| --- | --- | --- | --- | --- | --- |
| | General | HFS | Influenza vaccine | Psychophysical tests | Stimulated IL-6 and TNF-α |
| Not fluent in English | X |  |  |  |  |
| Incompetence to consent and participate, e.g. acute psychosis or high suicide risk | X |  |  |  |  |
| Pregnancy |  | X |  |  | X |
| Electrical implants (e.g. pacemaker) |  | X |  |  |  |
| Metal implants in the area receiving the HFS |  | X |  |  |  |
| Tattoos in the area receiving the HFS |  | X |  |  |  |
| Any visible injury or open wounds in the area receiving the HFS |  | X |  |  |  |
| Cardiovascular disorders |  | X |  |  |  |
| Known history of allergic reactions to vaccinations |  |  | X |  |  |
| Already received current season's influenza vaccination |  |  | X |  | X |
| Chronic pain (pain on most days for the past three months) |  |  |  | X | X |
| Diabetes mellitus |  | X |  |  | X |
| Peripheral vascular disease |  | X |  | X |  |
| Sensory impairment of areas to undergo psychophysical testing |  | X |  | X |  |
| Use of medication that could alter skin sensitivity (e.g. analgesic medication, topical medical creams in areas to undergo psychophysical testing) |  |  |  | X | X |
| Medication used to alter immune function (e.g. NSAIDs, steroids) |  |  |  |  | X |
| Smoking habit |  |  |  |  | X |
| Febrile illness in the past 4 weeks |  |  |  |  | X |

#### Screening and enrolment

To recruit participants with a varied range in childhood adversity, volunteers completed the 28-item Childhood Trauma Questionnaire-Short Form (CTQ-SF) [6]. Total CTQ-SF scores were used to categorise volunteers into three recruitment groups: (1) minimal (CTQ-SF score 25 – 36), (2) moderate (37 – 67), and (3) severe (> 67) childhood adversity [5]. We aimed to enrol 32 participants per childhood adversity group, enrolling on a ‘first to qualify and participate’ approach. Group allocation was used for recruitment purposes only and all participants, irrespective of group allocation, underwent the same procedure.

### Experimental manipulations

#### In vivo immune provocation

For the *in vivo* immune provocation, participants received the current season’s tetravalent influenza vaccine in the deltoid muscle of the test arm (i.e. the arm receiving the high-frequency electrical stimulation, contralateral to the arm used for the blood draw). Plasma IL-6 typically peaks approximately 24 hours after influenza vaccination [53]. Greater IL-6 expression at baseline is associated with increased pain at the vaccination site, body aches, and headaches after the influenza vaccine [10], linking IL-6 to nociceptive processing in this model.

#### In vitro immune provocation

The *in vitro* immune provocation required incubation of peripheral blood with lipopolysaccharide (LPS). Elevated expression of IL-6 to either *in vitro* [58] or *in vivo* [68, 69] LPS-provocation is associated with lower pressure pain threshold, linking cytokine responsiveness to nociceptive processing. On morning 1, peripheral blood was drawn into a TruCulture® tube pre-loaded with LPS and incubated at 37 DC for 24 hours. Thereafter, cells were separated from the supernatant and tubes were frozen at an initial –20 DC, followed by –80 DC for longer storage while awaiting batch analysis. All stimulated samples were assayed in duplicate (R&D 3-plex Discovery assay) at a dilution factor of 1:30, using Luminex xMAP technology, to estimate the levels of IL-1β, IL-6, and TNF-α. To estimate cytokine levels, we fitted a weighted quadratic model to define the standard curve and used the raw fluorescence values to interpolate estimates for samples that fell outside the assay’s expected range (details in Supplementary file: Section 3). In accordance with the study protocol, we report data for IL-6 and TNF-α, and our statistical analyses used a composite score of the mean of z-scores for IL-6 and TNF-α expression.

#### In vivo neural provocation

For the *in vivo* neural provocation, participants received high-frequency electrical stimulation (HFS) at one forearm. HFS was delivered using a constant current stimulation system (DS7A, Digitimer Limited, Hertfordshire, UK) to one pair of specialised surface electrodes on the test arm, as previously described [4]. HFS was delivered at ten times the current of the individual’s detection threshold, which was determined using an adaptive staircase method (see details in Supplementary file: Section 4). The HFS consisted of five one-second trains, using a two-millisecond pulse width of 100 Hz frequency, with a nine-second break between trains.

### Primary and secondary psychophysical outcomes (hypotheses 2 and 3)

Vulnerability to persistent pain was operationalised differently for each hypothesis, given the distinct experimental manipulations. For hypothesis 2, the HFS neural provocation largely targets spinal cord mechanisms; therefore, vulnerability to persistent pain was operationalised using static psychophysical tests of the (1) surface area (primary outcome) and (2) magnitude (secondary outcome) of HFS-induced secondary hyperalgesia to mechanical stimulation. For hypothesis 3, the influenza vaccine immune provocation typically has a systemic effect; therefore, vulnerability to persistent pain was operationalised using dynamic psychophysical tests of (1) conditioned pain modulation (primary outcome) and (2) temporal summation (secondary outcome). These different operationalisations aimed to provide broader phenotyping of each participant.

#### Primary outcome for hypothesis 2: surface area of mechanical secondary hyperalgesia

The surface area of secondary skin hyperalgesia (in cm^2^) was assessed using a 128 mN von Frey filament (MARSTOCK, Schriesheim, Germany), as described previously [4] (Fig 1), at 30, 45, and 50 minutes after the HFS induction. We included each participant’s three measures of surface area across each of the three time points in our statistical analysis (protocol deviation 1 of 4; Supplementary file: Section 2, Table S2).

**Figure 1:**
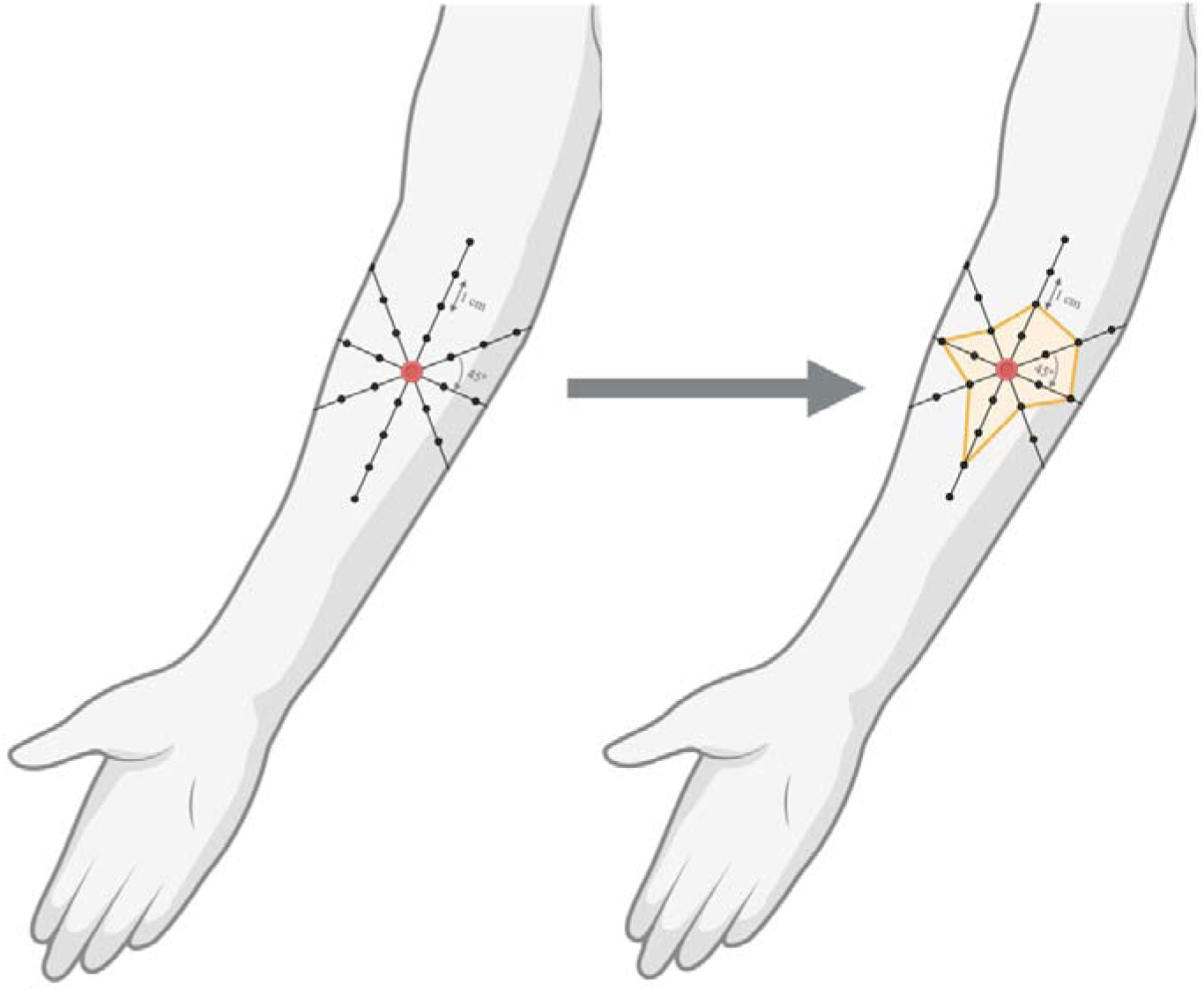
Eight-radial-lines approach to estimating surface area of secondary hyperalgesia. Viewing from left to right: An image of eight radial lines originating at the electrode, at 45D to each other (left). Dots along the lines are 1 centimetre apart and indicate the sites for test stimuli. An example of a mapped area of secondary hyperalgesia (right). The orange lines indicate the border of the estimated area of secondary hyperalgesia. Figure created in BioRender. Madden, T (2025) https://BioRender.com/q32i088.

#### Secondary outcome for hypothesis 2: magnitude of mechanical secondary hyperalgesia

The magnitude of secondary hyperalgesia to mechanical punctate stimulation was assessed adjacent to the electrode, using two punctate “pinprick” stimulators that exerted forces of 128 mN and 256 mN (MRS Systems, Heidelberg, Germany). Participants provided stimulus ratings using the Sensation and Pain Rating Scale (SPARS) (Fig 2) [41]. Ratings of a single set of these stimuli were taken before and 35, 50, and 65 minutes after the HFS induction. We included ratings for each stimuli at baseline and each of the three follow-up points for each participant in our statistical analysis (protocol deviation 2 of 4; Supplementary file: Section 2, Table S2).

**Figure 2:**
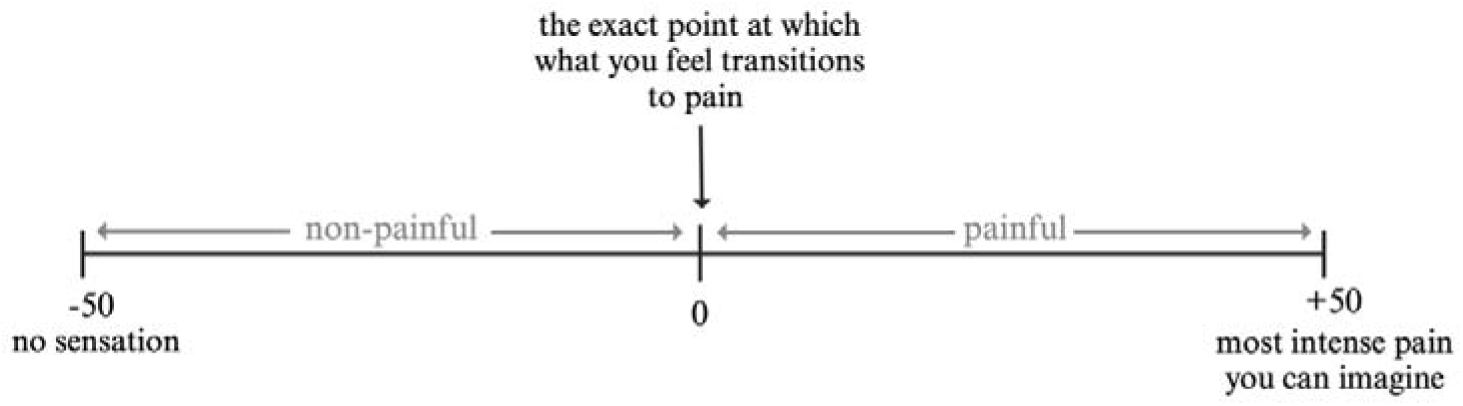
Sensation and Pain Rating Scale (SPARS) adapted from Madden, Kamerman [41]. On the left of the scale, the ‘non-painful’ range operates from –50 – “no sensation” to 0 – “the exact point at which what you feel transitions to pain”. On the right of the scale, the ‘painful’ range operates from 0 to +50 – “most intense pain you can imagine”. Figure created in BioRender. Madden, T. (2025) https://BioRender.com/j20o079.

#### Primary outcome for hypothesis 3: change in conditioned pain modulation

We estimated conditioned pain modulation (CPM) at the lumbar region (primary test site; in line with and 2 cm lateral to L2) to capture the systemic effect of the provoked immune response and at the deltoid insertion (secondary test site near the vaccination site), to capture the local effects of the provoked immune response. First, pressure pain threshold (test stimulus) was assessed with a hand-held algometer and a rate of change in pressure of ∼5 N per second until report of first pain. Second, the participant’s contralateral hand to the vaccination site was immersed in circulating cold water of ∼3 – 5 DC (conditioning stimulus). Third, when pain in the immersed hand reached +20 on the SPARS [41], pressure pain threshold was reassessed with the contralateral hand still immersed. Fourth, the hand was removed and wrapped in a towel for recovery. Fifth, when the participant reported that the previously immersed hand felt “normal again”, the pressure pain threshold was reassessed (results not reported here). This paradigm has excellent test-retest reliability in intra-session and 3-day test intervals [27]. CPM was estimated by subtracting the pressure pain threshold before immersion from the pressure pain threshold during cold water immersion. The dependent variable for hypothesis 3 was the change in CPM between mornings, i.e. CPM 24h *after* the influenza vaccine (i.e. morning 2) minus CPM *before* the influenza vaccine (i.e. morning 1), such that a negative score would represent less efficient modulation on morning 2 than on morning 1.

#### Secondary outcome for hypothesis 3: change in temporal summation

Temporal summation (TS) was assessed before CPM at both the lumbar and deltoid test sites by subtracting the SPARS rating of a single stimulation from the SPARS rating of the final of 16 stimulations at 60 Hz using a 256 mN Von Frey filament [1]. The dependent variable for hypothesis 3 was the change in TS between both mornings, i.e. TS 24h *after* the influenza vaccine (i.e. morning 2) minus TS *before* the influenza vaccine (i.e. morning 1), such that a positive score will represent more efficient summation on morning 2 than on morning 1.

### Exploratory outcomes

#### Static and dynamic light touch and single electrical stimulation

As exploratory outcomes to inform future studies, we also assessed SPARS ratings to static (32 mN von Frey filament [55]) and dynamic (soft brush [35]) light touch and single electrical stimulation (2 ms pulse duration; current 10x individual electrical detection threshold [21]) before and after the HFS induction, at the same time points as mechanical punctate stimulation.

### Potential confounding factors

Candidate confounders were prioritised for assessment: positive childhood experiences, long-term stress, depression and anxiety, asthma, COVID-19 infection, chronic and recent illnesses, and sleep (for details on the outcome measures for each potential confounding factor, see Supplementary file: Section 5). The process of selecting candidate confounders was guided by a four-pronged approach: we constructed a directed acyclic graph, consulted with experts in the field, thoroughly reviewed the literature, and evaluated the feasibility of assessing of potential confounders. For each candidate confounder, we tested for an association with the study outcome or relationship of interest. Total score on the CTQ-SF was also included as a potential confounder for hypotheses 2 and 3.

### Procedure

Figure 3 shows the study procedure. Blood drawn at the start of the procedure on morning 1 was used for the *in vitro* LPS immune provocation. The 24-hour period after the influenza vaccine was administered (on morning 1) coincides with the approximate peak immune response to the influenza vaccine [53]. The 24-hour circadian rhythmicity of endogenous cortisol is thought to influence variability in innate immune responses. In the early morning, cortisol levels are high and cytokine levels are low; in the late afternoon, cortisol levels are low and cytokine levels are high. Data on the relationship between circadian-driven changes in cortisol and LPS-provoked cytokines are conflicting, with some studies finding a negative association between endogenous cortisol and LPS-provoked cytokines [13, 50], and others finding no relationship [22]. To account for the potential influence of circadian rhythm-driven variability on innate immune responses, all testing sessions began before 12:00 noon [57, 67].

**Figure 3:**
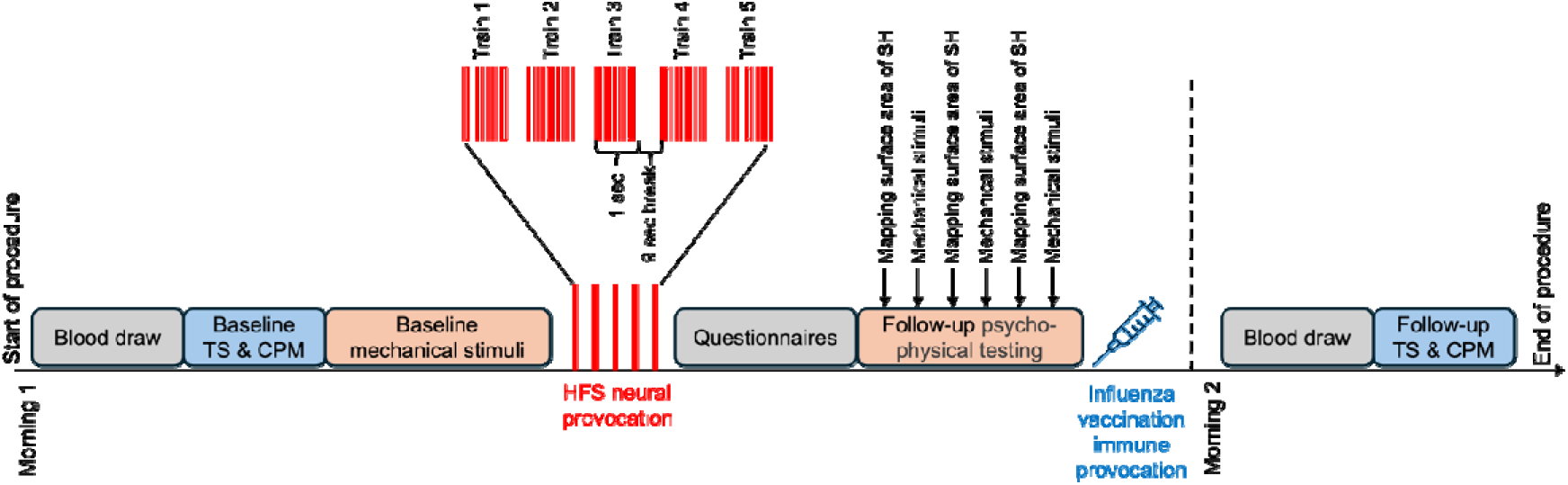
Study procedure. The first blood draw on Morning 1 was used for the in vitro LPS provocation. The second blood draw on Morning 2 is for another study, and results are not reported in this report. CPM: conditioned pain modulation; HFS – high-frequency electrical stimulation; SH: secondary hyperalgesia; TS: temporal summation.

#### Blinding of participants

Participants were blinded to the research questions and hypotheses of this study. The study information sheet merely informed participants that “we want to understand how early life experiences affect the immune and neural systems”. To assess if blinding was maintained, participants were asked at the end of the procedure to explain what they thought the purpose of the study was. The assessor (GJB) judged if blinding was maintained or broken based on the participant’s response, using conservative criteria – i.e. leaning towards confirming unblinding if given any hint of that possibility. Broken blinding is reported descriptively, and sensitivity analyses were conducted to investigate the influence of broken blinding on the study results.

#### Blinding of the assessor

The assessor (GJB) was blinded to each participant’s childhood adversity group allocation but not to the study aims. After each testing procedure, the assessor completed a blinding assessment for each participant, for which the assessor stated (or guessed) in which group (mild, moderate, or severe childhood adversity) each participant belonged and rated their confidence on a Likert scale (“not at all confident”, “not confident”, “I don’t know”, “confident”, “extremely confident”). Broken blinding was assessed using the chi-squared test (protocol deviation 3 of 4; Supplementary file: Section 2, Table S2) and reported descriptively, and sensitivity analyses were conducted to investigate the influence of broken blinding on the study results.

### Statistical analysis

#### Sample size calculations

The target sample size needed to balance pragmatism with adequate power. In the absence of suitable pilot data to inform a sample size calculation and the unavailability of methods to calculate sample size to support all three hypotheses, we estimated the sample size that would provide reasonable power for each hypothesis with alpha 0.05, power 0.8, and used the largest estimate of the three, which was n = 96. Therefore, we aimed for complete datasets from 96 participants.

After data collection and before finalising the R analysis script, we recognised an error in interpreting the sample size calculations: the target sample size should have been 85 for a correlation coefficient of 0.3. However, we wished to use the data we had collected from the full sample of 96 participants. Therefore, we used G*Power [16] to conduct a sensitivity power analysis (Supplementary file: Section 6, Fig S1), which estimated that our final sample size (n=96) provided *a priori* power to detect an effect size of r = 0.275 with power 0.8 and alpha 0.05. This calculation was completed before the actual study data were processed.

#### Statistical analysis plan

Before the formal data were analysed, the study protocol and pilot data analysis script were registered and locked on the Open Science Framework’s online platform *[link provided at publication]*. For all three research questions, we followed best practice by using both visual data analysis and formal modelling to investigate the relationships specified in the three hypotheses. The specifics of the models were determined by the data features to achieve the best-fitting model that is interpretable. Data were analysed using R (version 4.4.0, packages: readr [74], tidyverse [72], magrittr [44], ggplot2 [70], dplyr [73], lmtest [76], lmerTest [33], brms [9], emmeans [36], tidybayes [26], broom [54], broom.mixed [7], scales [71], patchwork [49], sjPlot [39]) in RStudio [56].

#### Assessment of model fit

An assessment of model fit was conducted for the unadjusted and covariate-adjusted models. Four assumptions were assessed: (1) linearity, (2) homoscedasticity, (3) normally distributed residuals, and (4) no influential observations. The model was deemed unfit for these data if any assumptions were violated.

#### Manipulation checks

For hypothesis 3, we conducted two manipulation checks. First, a statistically significant difference in pressure pain threshold before, compared to during the cold water immersion indicated a successful CPM procedure. Second, for TS, a statistically significant difference in SPARS ratings to a single stimulus compared to the 16th stimulus indicated a successful TS procedure.

## Results

### Participants

A total of 101 participants were enrolled and tested in this study. Five participants’ data were excluded from the formal data analysis (n = 3 data were not saved due to technical issues; n = 1 did not complete testing (morning 2); n = 1 disclosed a smoking habit after the procedure) (Fig 4). Therefore, data from 96 participants (61 females; median (range) age: 23 (18 – 65) years old) were included in the formal data analysis. There were complete datasets for all outcomes except for TS at the lumbar site, for which data were missing for one participant due to a technical issue. This participant was excluded only from the analysis of TS at the lumbar site. A summary of the descriptive statistics are presented in Table 2 and Supplementary file: Section 7, Table S3.

**Figure 4:**
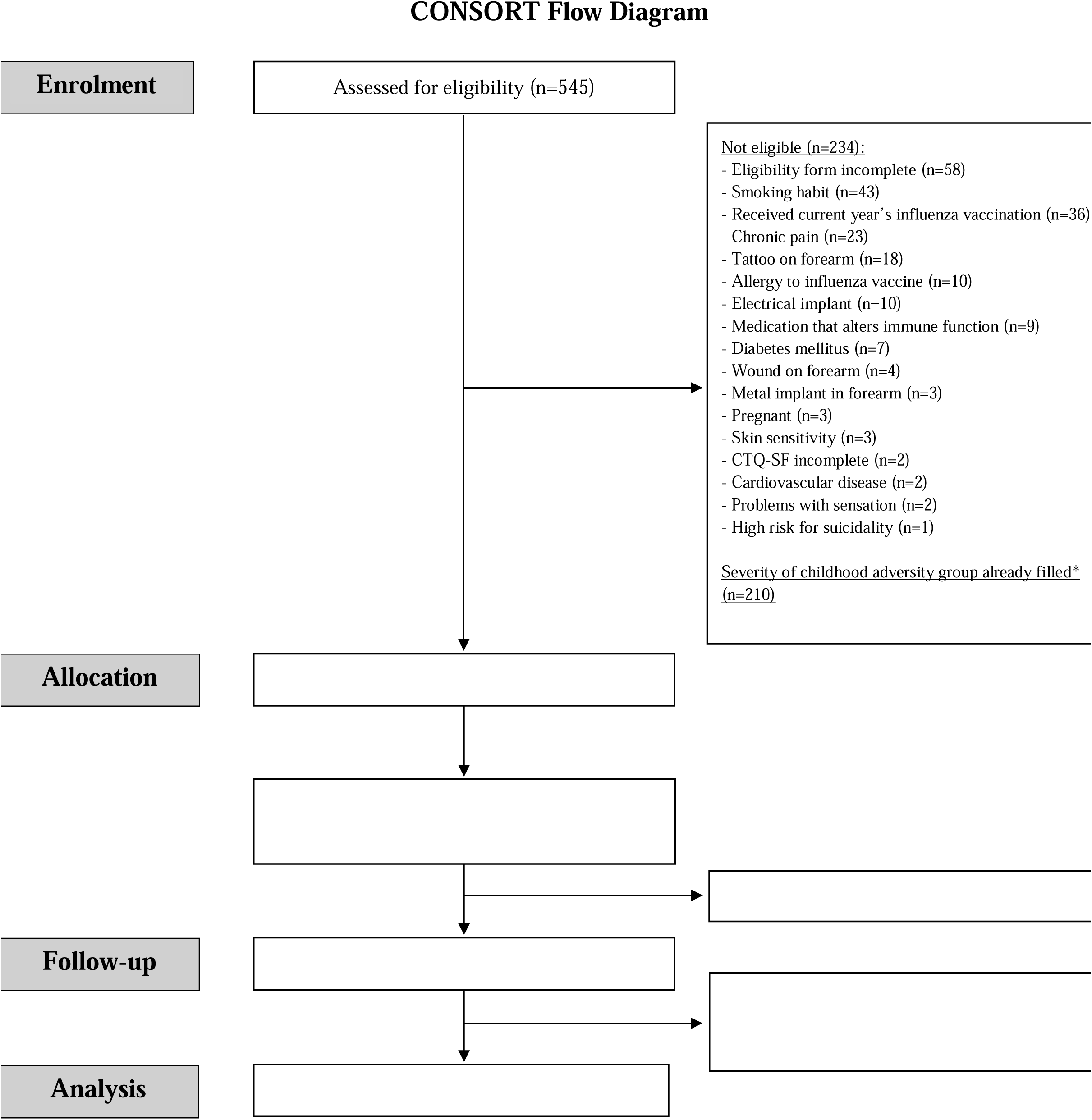
CONSORT flow diagram. *We aimed to enrol 32 participants per childhood adversity group, enrolling on a ‘first to qualify and participate’ approach. See *Screening and enrolment* for more details. CTQ-SF: Childhood Trauma Questionnaire-Short Form.

**Table 2:** Descriptive statistics of participants’ characteristics.

| Characteristics | Full sample<br>N = 96 | History of mild childhood adversity (CTQ-SF score: 25 – 36)<br>N = 32 | History of moderate childhood adversity (CTQ-SF score: 37 – 67)<br>N = 32 | History of severe childhood adversity (CTQ-SF score: 68 – 125)<br>N = 32 | p-value for between-group differences |
| --- | --- | --- | --- | --- | --- |
| Age (years) | 23 (21– 30) | 23 (21 – 27) | 22 (21 – 25) | 24 (22 – 35) | 0.21 |
| Sex (n male:female) | 35:61 | 8:24 | 20:12 | 7:25 | < 0.001<sup>s</sup> |
| **Cytokines (pg/mL)** |  |  |  |  |  |
| Provoked IL-6 | 31331 (20947.9 – 31644.3) | 31331 (20992.5 – 31649.7) | 22502.3 (17268.2 – 31649.7) | 31642.5 (31331.0 – 31642.5) | 0.02 |
| Provoked TNF-α | 4032.8 (2884.1 – 5402.0) | 3967 (3044.4 – 5403.4) | 3577.7 (2767.5 – 4749.5) | 4411.3 (3423.3 – 6284.9) | 0.34 |
| Current used for HFS (mA) | 0.17 (0.13 – 0.23) | 0.17 (0.16 – 0.21) | 0.20 (0.17 – 0.27) | 0.13 (0.12 – 0.23) | 0.001 |
| **Surface area of secondary hyperalgesia (cm<sup>2</sup>)** |  |  |  |  |  |
| 30 min after HFS | 22.19 (6.19 – 37.60) | 10.80 (3.93 – 28.08) | 22.97 (12.37 – 32.01) | 31.22 (3.04 – 47.32) | 0.08 |
| 45 min after HFS | 17.28 (8.05 – 36.13) | 15.71 (5.50 – 28.96) | 19.83 (12.37 – 35.54) | 18.85 (7.66 – 42.51) | 0.22 |
| 60 min after HFS | 17.5 (3.83 – 31.90) | 14.92 (3.93 – 26.02) | 17.48 (3.63 – 32.50) | 20.03 (2.16 – 39.76) | 0.129 |
| **Ratings to mechanical punctate stimulation (SPARS)** |  |  |  |  |  |
| Before HFS | -21.81 (-34.35 – 1.27) | -20.91 (-32.36 – 2.31) | -18.72 (-31.93 – 2.26) | -25.13 (-39.25 – 0.44) | 0.53 |
| 35 min after HFS | -12.10 (-30.88 – 3.03) | -18.71 (-24.71 – 4.22) | -11.29 (-26.63 – 4.79) | -9.35 (-36.16 – 0.15) | 0.53 |
| 50 min after HFS | -9.62 (-25.97 – 3.03) | -10.64 (-22.67 – 8.27) | -7.94 (-26.14 – 3.62) | -7.46 (-26.06 – 2.94) | 0.92 |
| 65 min after HFS | -7.86 (-26.78 – 5.61) | -7.86 (-22.58 – 8.69) | -2.34 (-28.65 – 5.46) | -9.80 (-33.37 – 3.21) | 0.77 |
| **Conditioned pain modulation** (change in pressure pain threshold, N) |  |  |  |  |  |
| <u>Lumbar test site</u> |  |  |  |  |  |
| Before immune provocation | 20.8 (12.50 – 29.90) | 19.35 (14.64 – 22.94) | 22.33 (11.86 – 30.64) | 19.98 (8.18 – 32.60) | 0.80 |
| After immune provocation | 18.8 (7.98 – 27.70) | 15.23 (6.78 – 28.60) | 22.80 (12.08 – 33.25) | 17.88 (4.70 – 22.05) | 0.07 |
| <u>Deltoid test site</u> |  |  |  |  |  |
| Before immune provocation | 11.53 (5.94 – 18.76) | 6.78 (4.93 – 13.81) | 14.33 (9.49 – 22.25) | 13.38 (5.19 – 19.60) | **0.01** |
| After immune provocation | 11.68 (6.03 – 17.16) | 7.28 (5.33 – 12.65) | 13.53 (10.38 – 24.20) | 12.00 (5.49 – 17.70) | **0.01** |
| **Temporal summation** (change in SPARS ratings, 16<sup>th</sup> minus 1st) |  |  |  |  |  |
| <u>Lumbar test site</u> |  |  |  |  |  |
| Before immune provocation | 6.19 (1.27 – 19.25) | 7.22 (1.51 – 17.82) | 5.87 (1.35 – 16.51) | 5.00 (0.48 – 21.29) | 0.24 |
| After immune provocation | 4.64 (1.11 – 11.20) | 7.86 (3.06 – 21.65) | 6.15 (1.401 – 10.69) | 1.79 (0.32 – 6.94) | **0.01** |
| <u>Deltoid test site</u> |  |  |  |  |  |
| Before immune provocation | 7.50 (1.05 – 17.96) | 8.06 (2.44 – 13.45) | 8.06 (1.036 – 18.10) | 5.95 (0.77 – 19.98) | 0.78 |
| After immune provocation | 3.89 (-0.02 – 10.10) | 4.92 (1.35 – 10.18) | 2.86 (-0.91 – 6.98) | 3.02 (-0.36 – 12.46) | 0.68 |
| **Adverse childhood experiences (CTQ-SF)\*** | 49.0 (33.0 – 74.0) | 30.5 (27.8 – 33.0) | 49.0 (44.5 – 56.0) | 83.0 (74.0 – 93.0) | **< 0.001** |
| Subscale: physical abuse<sup>†</sup> | 8.0 (5.0 – 14.3) | 5.0 (5.0 – 6.0) | 7.5 (6.0 – 10.0) | 16.0 (13.0 – 20.3) | **< 0.001** |
| Subscale: emotional abuse<sup>†</sup> | 10.5 (7.0 – 19.0) | 6.5 (5.0 – 8.0) | 10.0 (9 – 13.5) | 21.0 (19.0 – 22.0) | **< 0.001** |
| Subscale: sexual abuse<sup>†</sup> | 5.0 (5.0 – 13.0) | 5.0 (5.0 – 5.0) | 5.0 (5.0 – 9.5) | 14.0 (6.8 – 20.3) | **< 0.001** |
| Subscale: physical neglect<sup>†</sup> | 8.0 (5.0 – 13.0) | 5.0 (5.0 – 6.0) | 9.0 (6.0 – 10.3) | 14 (10.8 – 17.0) | < **0.001** |
| Subscale: emotional neglect<sup>†</sup> | 13.0 (7.0 – 19.0) | 6.0 (5.0 – 8.3) | 13.0 (9.0 – 15.3) | 20.0 (18.0 – 21.3) | < **0.001** |

CTQ-SF = Childhood Trauma Questionnaire-short form; SPARS = Sensation and Pain Rating Scale; HFS = high-frequency electrical stimulation.
\* Possible total score range for the CTQ-SF: 25 – 125.
<sup>†</sup> Possible total score range for each subscale of the CTQ-SF: 5 – 25.
<sup>§</sup> Statistical test used for sex was a Pearson's Chi-squared test. Statistical test for all other variables was an ANOVA.
Data are presented as median (IQR), mean ( ± SD), or n (%)

### Manipulation checks

#### Pressure pain threshold and conditioned pain modulation (CPM)

On average, CPM was successfully induced at the sample level at both test sites (deltoid and lumbar) and at both test sessions (before and after the influenza vaccine): Pressure pain threshold was higher during the cold water immersion than before, at both the lumbar and deltoid test sites (Fig 5). On average, the cold water conditioning stimulus increased pressure pain threshold by 20.49 N [95% CI: 17.80;23.17; *p* <0.001] at the lumbar site and 13.08 N [95% CI: 1.37;14.80; *p* <0.001] at the deltoid site (Fig 5, and Supplementary file: Section 8, Table S4).

**Figure 5:**
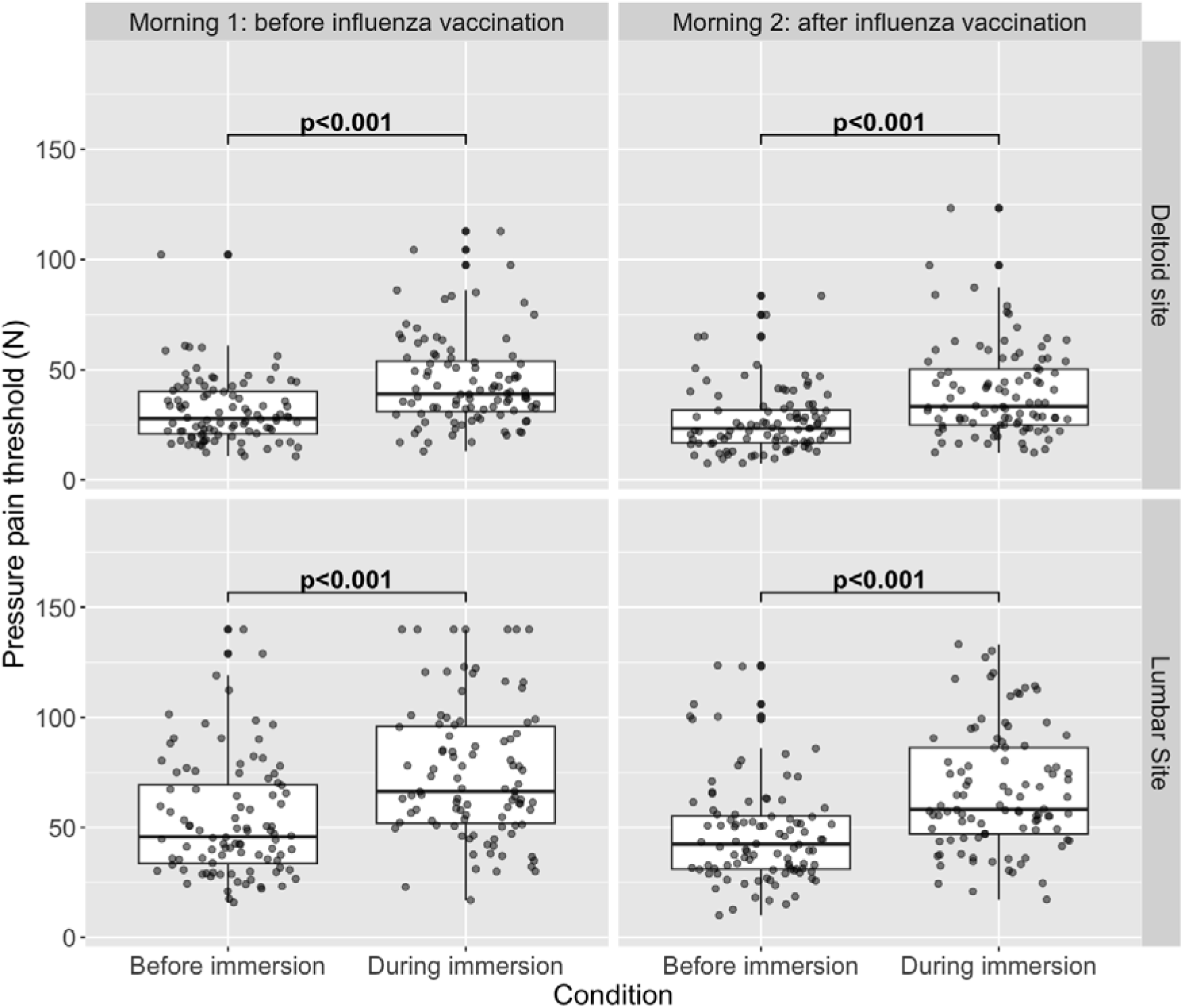
Boxplots of pressure pain threshold before and during cold water immersion, faceted by session (i.e. morning 1 and 2) and test site.

#### SPARS rating to mechanical stimuli and temporal summation (TS)

On average, TS was successfully elicited at the sample level at both test sites (deltoid and lumbar) and at both test sessions (before and after the influenza vaccine): SPARS ratings were higher to the 16^th^ of the 16 stimuli than to the single mechanical stimulus at both the lumbar and deltoid sites (Fig 6). On average, there was a 9.50 [95% CI: 6.99;12.00; *p* <0.001] unit increase in SPARS rating at the lumbar site and an 8.57 unit [95% CI: 6.26;10.88; *p* <0.001] increase in SPARS rating at the deltoid site to the 16^th^ mechanical stimulation (Fig 6, and Supplementary file: Section 8, Table S5). Therefore, TS was successfully induced at the sample level at both test sites (deltoid and lumbar) and at both test sessions (before and after the influenza vaccine).

**Figure 6:**
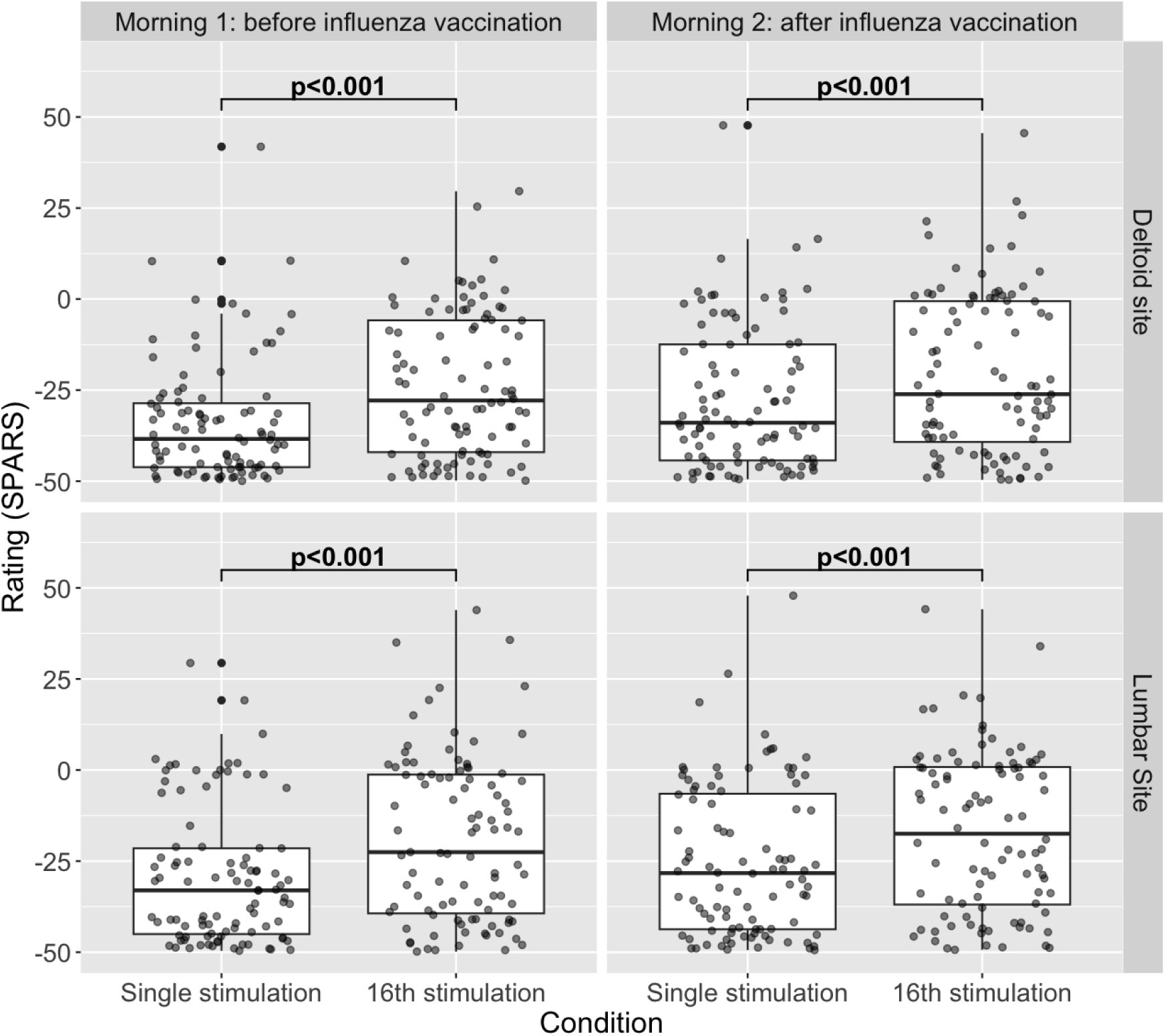
Boxplots of ratings to single and 16^th^ mechanical stimulation, faceted by session (i.e. morning 1 and 2) and test site.

### Hypothesis 1: relationship between childhood adversity and provoked cytokine expression

We tested whether the CTQ-SF total score was positively associated with provoked cytokine expression using simple linear regression. Both unadjusted and covariate-adjusted models satisfied the underlying assumptions of linear regression (Supplementary file: Section 9, Fig S2). Neither model found that CTQ-SF total score was associated with cytokine expression (*p-values =* 0.182 and 0.092; Fig 7 and Supplementary file: Section 9, Table S6).

**Figure 7:**
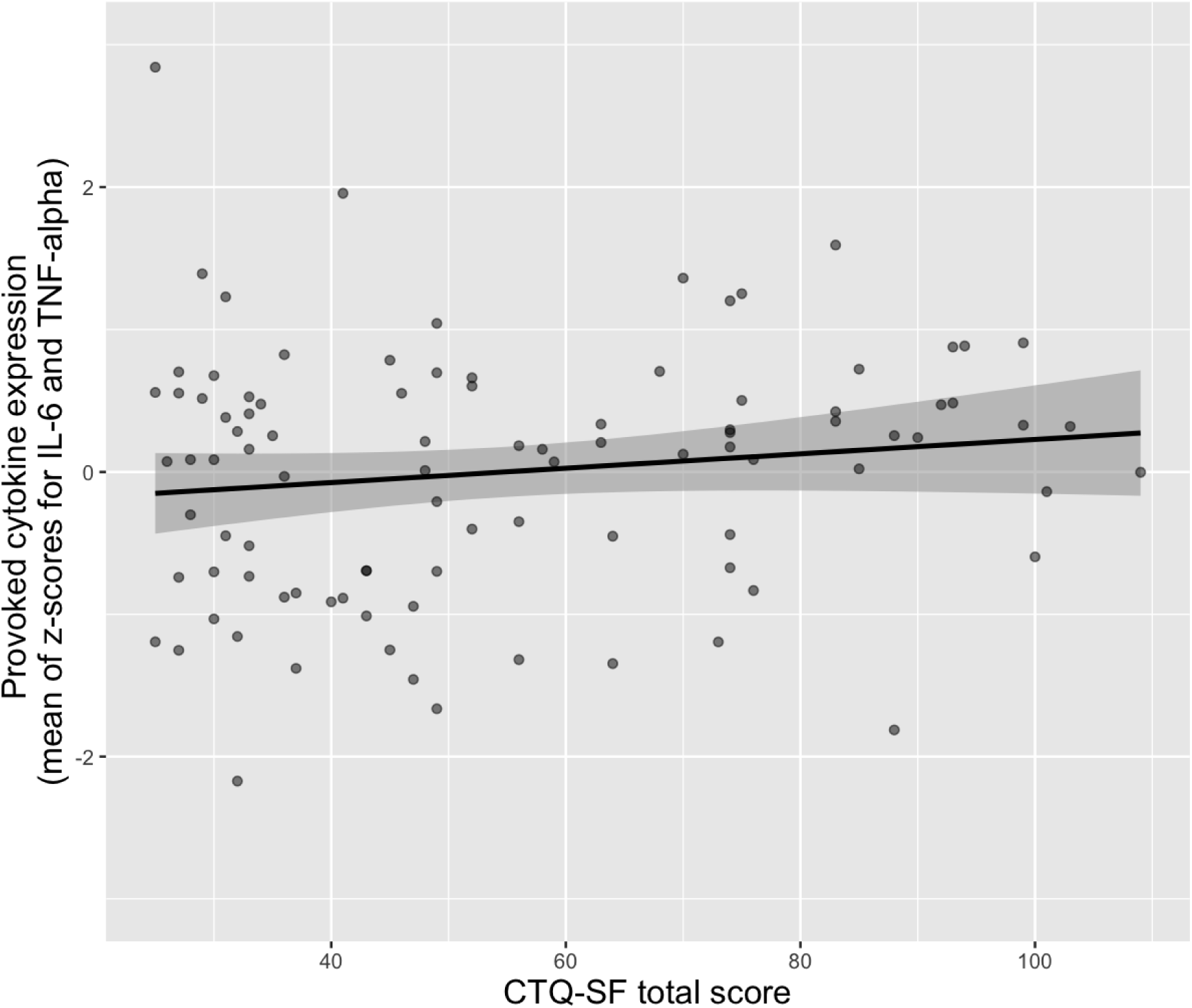
Relationship between CTQ-SF score and provoked cytokine expression (n = 96). CTQ-SF: Childhood Trauma Questionnaire-Short Form.

### Hypothesis 2: relationship between provoked cytokine expression and induced secondary hyperalgesia

We tested whether provoked cytokine expression was positively associated with the surface area (primary outcome) and magnitude (secondary outcome) of secondary hyperalgesia.

#### Primary analysis: surface area of secondary hyperalgesia

Conventional and robust regression modelling approaches violated the underlying assumptions of linear regression, showing noteworthy heterogeneity of variance (Supplementary file: Section 10, Figs S3 & S4), likely due to the high number of zero values (∼14%) for the outcome (i.e. no area of secondary hyperalgesia). Hurdle models are designed for data with many zero values and no upper bound. They incorporate two separate components: a conditional linear regression that models non-zero outcome data only and a logistic regression that assesses the value of the designated independent variables in predicting zero values. Provoked cytokine expression was not associated with surface area. This was true for the conditional (non-zero) and logistic regression portions of both unadjusted and covariate-adjusted hurdle models (Supplementary file: Section 10, Fig S5, Table S7).

#### Secondary analysis: magnitude of secondary hyperalgesia

The unadjusted and covariate-adjusted models satisfied the underlying assumptions of linear regression (Supplementary file: Section 10, Fig S6). Neither model found that provoked cytokine expression was associated with the magnitude of secondary hyperalgesia (*p-values =* 0.94 and 0.77; Supplementary file: Section 10, Fig S7 and Table S8).

### Hypothesis 3: relationship between provoked cytokine expression and change in CPM and TS

#### Primary analysis: change in CPM at the lumbar and deltoid test sites

We tested whether provoked cytokine expression was negatively associated with a change in CPM at the lumbar (primary test site) and the deltoid (secondary test site). CPM was no different before vs after the influenza vaccination, at either the lumbar (*p =* 0.32) or the deltoid site (*p* = 0.76; Supplementary file: Section 11, Table S9).

The unadjusted and covariate-adjusted models satisfied the underlying assumptions of linear regression (Supplementary file: Section 11, Figs S8 and S9). Neither model found a that provoked cytokine expression was associated with the change in CPM at the lumbar (*p-values =* 0.08 and 0.07, Fig 8A and Supplementary file: Section 11, Table S10) or at the deltoid test site (*p-values =* 0.27 and 0.33; Fig 8B and Supplementary file: Section 11, Table S11).

**Figure 8:**
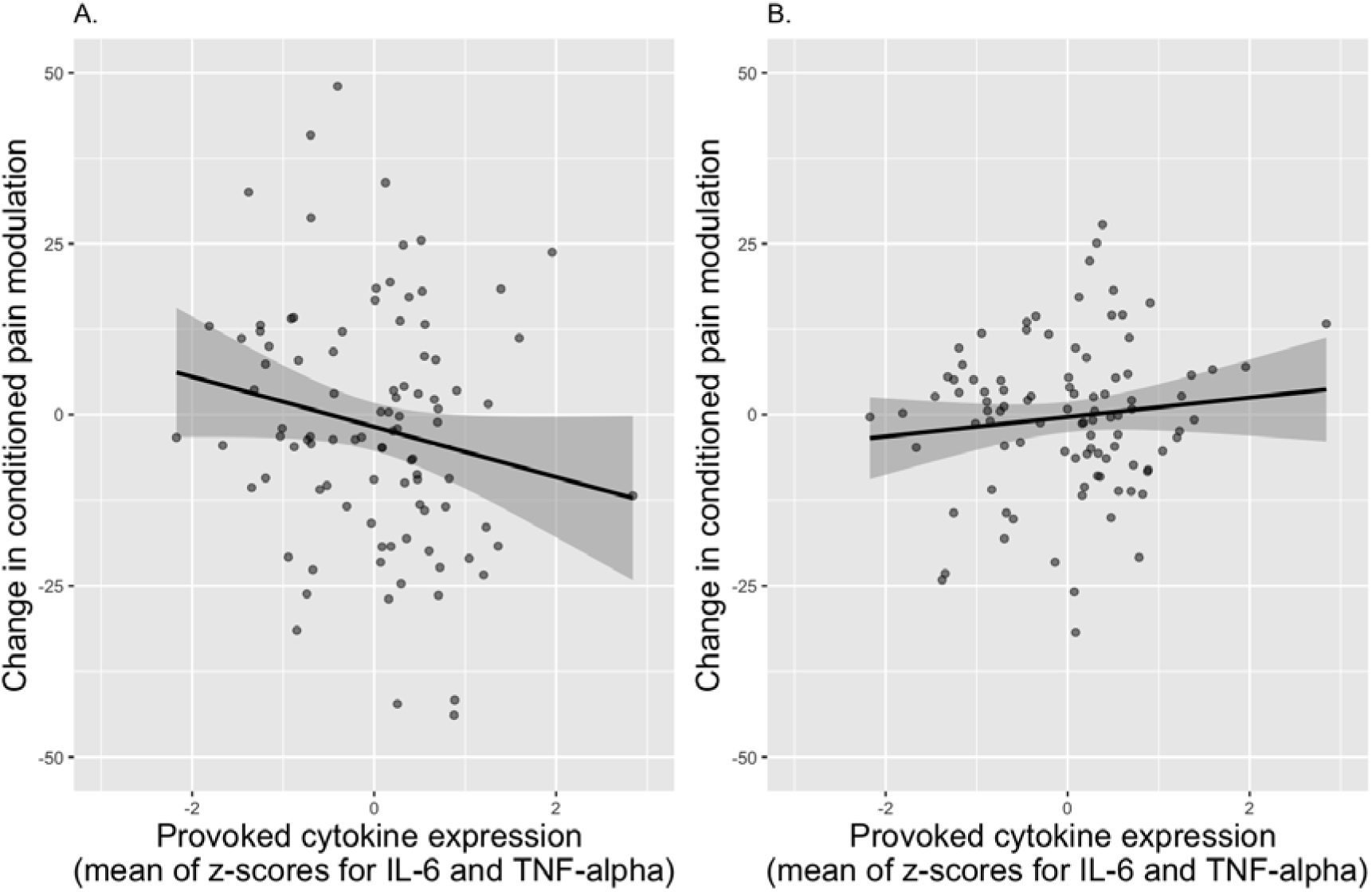
The relationship between provoked cytokine expression and change in conditioned pain modulation after immune provocation (influenza vaccination) at the lumbar test site. (A) (n=96) and deltoid test site (B) (n=96).

#### Stratifying by sex

Data suggest that females, but not males, show reduced conditioned pain modulation to an *in vivo* LPS challenge [25]. Consequently, we conducted an *ad hoc* exploratory analysis that stratified the relationship between our *in vitro* LPS-provoked cytokine expression and change in CPM at the lumbar and deltoid test sites. We observed no evidence of an association between provoked cytokine expression and change in CPM in either males or females at both test sites (Supplementary file: Section 11: Fig S10.)

#### Secondary analysis: change in TS at the lumbar and deltoid test sites

We tested whether provoked cytokine expression was positively associated with a change in TS. TS decreased from before to after the influenza vaccination at the deltoid site (*p* = 0.02) but not at the lumbar site (*p =* 0.09). On average, the influenza vaccine reduced TS by 3.19 [95% CI: –5.96; –0.42] units at the deltoid site (Supplementary file: Section 11, Table S12). Therefore, TS was successfully altered by the *in vivo* immune provocation (i.e. influenza vaccine) only at the deltoid site at the sample level.

The unadjusted and covariate-adjusted models satisfied the underlying assumptions of linear regression (Supplementary file: Section 11, Figs S11 and S12). Neither model found that provoked cytokine expression was associated with the change in TS at the lumbar (*p-values =* 1.0 and 0.92; Fig 9A and Supplementary file: Section 10, Table S13) or at the deltoid test site (*p-values =* 0.80 and 0.66; Fig 9B and Supplementary file: Section 11, Table S14).

**Figure 9:**
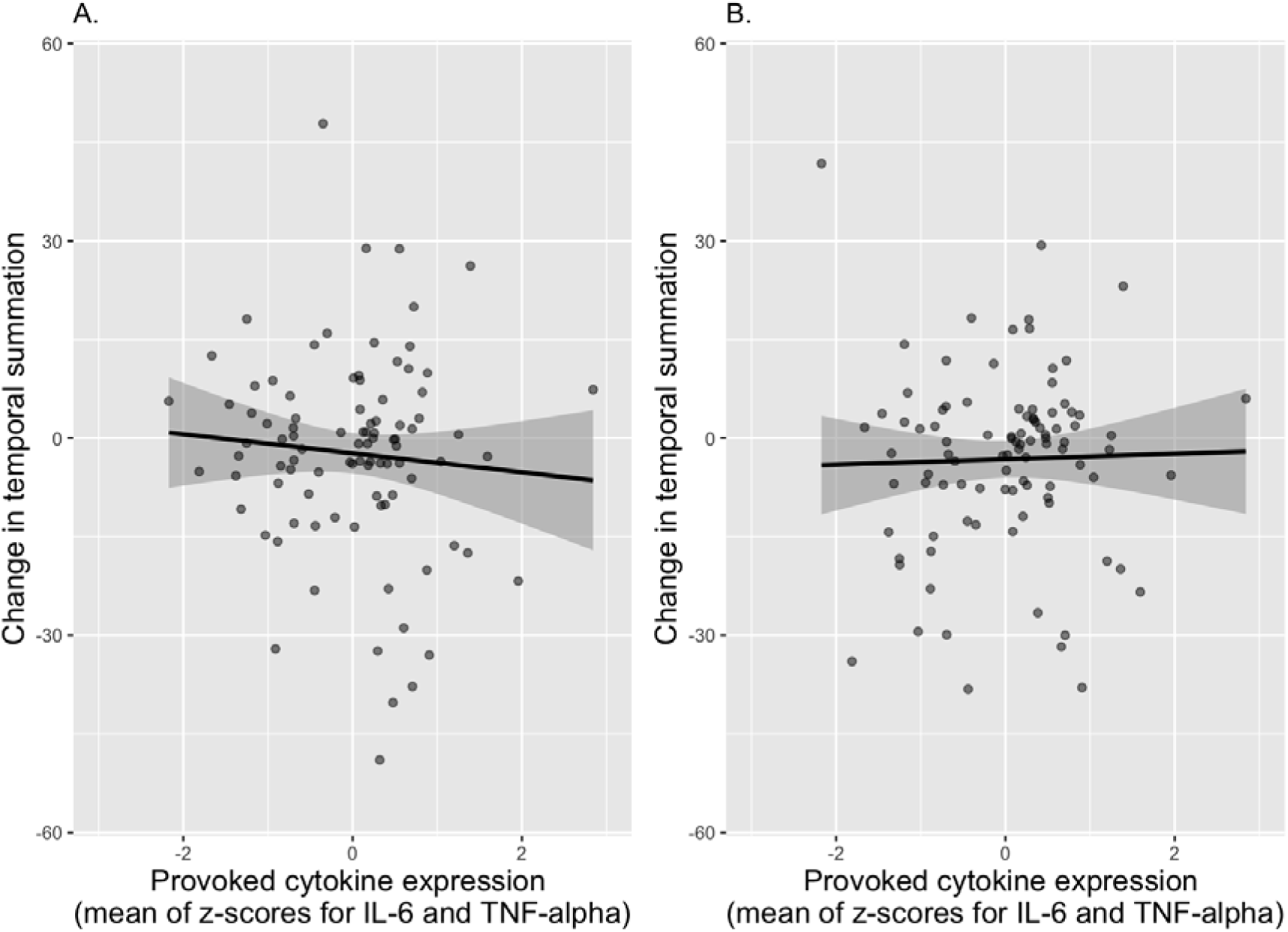
The relationship between provoked cytokine expression and change in temporal summation after the immune provocation (influenza vaccination) at the lumbar test site. (A) (n= 95) and deltoid test site (B) (n=96).

### Blinding assessments

#### Blinding of participants

Six (of 96) participants were unblinded to one of the 3 hypotheses, n = 2 for hypothesis 1 and n = 4 for hypothesis 3. No participant was unblinded to hypothesis 2. Sensitivity analyses were conducted for hypotheses 1 and 3, excluding unblinded participants. They showed no noteworthy changes in the association between CTQ-SF total score and provoked cytokine expression (hypothesis 1) (Supplementary file: Section 12, Table S15) nor between provoked cytokine expression and change in CPM or TS (hypothesis 3) (Supplementary file: Section 12, Tables S16 – S19).

#### Blinding of the assessor

Data on the assessor’s guess of group allocation were missing for one participant (of 96). The assessor correctly guessed group allocation for 44 participants (46.3% of n=95;). Visualisation suggested no relationship between guess accuracy and guess confidence (Supplementary file, Section 12, Fig S13), but a chi-square test showed a statistically significant difference (*p-value* = 0.011) between the assessor’s guessed group allocation and the actual group allocation, indicating the assessor’s guesses of group allocation were not random (as would be seen if blinding was maintained); therefore, blinding may have been broken. The planned sensitivity analysis was deemed unnecessary, given the lack of association between LPS-provoked cytokines and total score on the CTQ-SF.

### Exploratory analyses

#### Relationship between provoked cytokine expression and static and dynamic light touch and single electrical stimulation

Both the unadjusted and covariate-adjusted models found no main effect of provoked cytokine expression on static light touch (*p-values* = 0.45 and 0.48), dynamic light touch (*p-values* = 0.35 and 0.22), and single electrical stimulation (*p-values* = 0.46 and 0.29) (Supplementary file: Section 13, Table S20).

#### Relationship between each subscale of the CTQ-SF and provoked cytokine expression

We conducted an exploratory post-hoc analysis on the association between each subscale of the CTQ-SF and provoked cytokine expression. The sexual abuse subscale of the CTQ-SF was weakly correlated with provoked cytokine expression (r = 0.21, 95%CI: 0.01;0.4, *p =* 0.037) (Supplementary file: Section 13, Fig S14). None of the four other subscales of the CTQ-SF were correlated with provoked cytokine expression.

#### Interaction between positive childhood experiences and adverse childhood experiences on provoked cytokine expression

Data were available on positive childhood experiences (using total score from the Positive Childhood Experiences Questionnaire) for 49 (of 96) participants. Given the possibility that positive childhood experiences may moderate the influence of childhood adversity on the inflammatory response, we used these data to explore for an effect of the interaction between positive childhood experiences and total CTQ-SF score (i.e. adverse childhood experiences) on provoked cytokine expression. The interaction term was not statistically significant (*p* = 0.73), and the main effect of the CTQ-SF score remained statistically insignificant (*p* = 0.36) for this subsample of 49 participants (Supplementary file: Section 13, Table S21).

## Discussion

This study aimed to take the first steps towards clarifying neural and immune reactivity as a mechanistic link between childhood adversity and nociceptive processing. In a two-day experiment, we successfully induced secondary hyperalgesia, CPM, and TS and used an influenza vaccine to manipulate pain-related psychophysical outcomes. None of the hypotheses was upheld: LPS-provoked *in vitro* expression of pro-inflammatory cytokines was not related to childhood adversity (hypothesis 1), nor to induced secondary hyperalgesia (hypothesis 2), nor to vaccine-associated change in CPM or TS (hypothesis 3).

Childhood adversity has been consistently linked to elevated expression of *resting* pro-inflammatory cytokines. However, its association with *LPS-provoked* pro-inflammatory cytokines is more controversial. Meta-analytical synthesis of 25 studies estimated a significant, although small, association between childhood adversity and elevated resting expression of IL-6 and TNF-α in healthy adults [3]. The few studies that have investigated the relationship between childhood adversity and *LPS-provoked* pro-inflammatory cytokine expression present conflicting results. Converse to our results, in two different adult cohorts, total score on the CTQ-SF was associated with elevated expression of LPS-provoked IL-6 but not TNF-α [12, 28]. Notably, these cohorts included adults with or without current symptoms of depression or anxiety or a diagnosis of schizophrenia or schizoaffective disorder. Conversely, in adults institutionalised during their first year of life – an assumed adverse childhood event – no association was found between institutionalisation and either LPS-provoked IL-6 or TNF-α [15]. Adults who were separated from their biological parents during their first years of life presented with *lower* levels of LPS-provoked IL-6 than controls raised by their biological parents [14]. This discrepancy in the relationship between childhood adversity and resting versus provoked cytokine expression may be because LPS-provocation of pro-inflammatory cytokines provides insight into the propensity of the immune system to mount a response (i.e. immune reactivity), which is distinctly different from the resting state of the immune system.

An additional stressor may be needed to unmask an influence of childhood adversity on cytokine expression. Two studies found that adversities in childhood alone did not predict elevated expression of LPS-provoked cytokines; however, childhood adversities coupled with recent stress did predict the elevated expression of LPS-provoked cytokines [24, 40]. These results highlight the layering of multiple challenges to reveal an underlying phenotype.

The consistent positive association between childhood adversity and resting pro-inflammatory cytokine expression suggest that childhood adversity may have long-lasting effects on tonic immune activity. On the other hand, that childhood adversity is associated with provoked pro-inflammatory cytokine expression only in the presence of recent stress suggests that a childhood adversity may not have a long-lasting effect on provoked immune activity, and a recent challenge (e.g. recent stress) may have short-term effects on phasic immune activity. However, the relative importance of tonic versus phasic immune activity to meaningful clinical outcomes remains unknown.

Individuals with chronic pain exhibit elevated resting pro-inflammatory cytokines. This relationship suggests that immune reactivity may support hyperresponsiveness of nociceptive processing, thus indirectly contributing to the persistence of pain. However, this study’s systematic deconstruction of immune reactivity and spinal nociceptive reactivity in humans calls this idea into question. These conflicting findings must be held in balance with previous work in which *in vivo* LPS-provoked cytokines were associated with the surface area of capsaicin-induced hyperalgesia and allodynia in humans [23]. *In vivo* LPS may be more potent than *in vitro* LPS: the live system contains more cells to scale both direct and indirect responses to provocation than a 1mL blood sample, and active blood circulation likely enhances the reach of signalling proteins to target cellular interactions to increase responsiveness in a way that cannot be achieved during standing tube incubation.

Additionally, an immune provocation coupled with a neural provocation, rather than an immune provocation alone, may be required to sufficiently challenge the nociceptive system [68]. Hutchinson, Buijs [23] found that *in vivo* LPS-provoked cytokines were not associated with hyperalgesia and allodynia; however, after administration of a capsaicin neural provocation, *in vivo* LPS-provoked cytokines were associated with capsaicin-induced hyperalgesia and allodynia, suggesting that systemic inflammation exacerbates capsaicin-induced hyperalgesia. Our study electrically induced secondary hyperalgesia in an immune-unchallenged system and found no association between secondary hyperalgesia and *in vitro* LPS-provoked pro-inflammatory cytokines. However, had we induced secondary hyperalgesia *after* administering the *in vivo* immune provocation, i.e. influenza vaccination, induced secondary hyperalgesia may have been associated with vaccine-associated elevated expression of pro-inflammatory cytokines. Although the influence of systemic inflammation on HFS-induced secondary hyperalgesia is unknown, given administration of intradermal capsaicin induces hyperalgesia and allodynia that are thought to reflect the heterotopic long-term potentiation-like processes that is also seen with the HFS induction model, it is likely that systemic inflammation would also exacerbated HFS-induced hyperalgesia.

### Strengths

The study’s sample presents genetic and environmental features that differ from the features of samples that are more typical in heterogeneous psychoneuroimmunology studies. Systematic reviews on the relationships between childhood adversity, pain, and immune reactivity typically involve homogenous samples from high-income countries with similar genetic and environmental factors. When drawing inferences about fundamental principles of psychoneuroimmunology, leaning into a literature that draws on a small slice of the human population runs the risk of biased conclusions. This is particularly important in light of genetic variability and environmental determinants in immune function: African ancestry is associated with larger immune variability and more pro-inflammatory phenotypes than European ancestry [38, 45, 52], and immune functioning is constantly shaped by environmental microbiota [38]. Our sample included participants with a variety of ancestries, including African, European, and South Asian; therefore, this study lays the foundation for future research to unpack the influence of genetic variability on immune reactivity in response to childhood adversity. We argue that there is an urgent need to correct the current dearth of immune-phenotyping and psychoneuroimmunology studies in low– and middle-income countries [38].

In addition to the strength of this study’s diverse sample, this study upheld the principles of open science: the protocol was registered at clinicaltrials.gov and locked online at Open Science Framework, all protocol deviations were declared, and de-identified data are available at *[insert GitHub link at publication]*.

### Limitations

Although the influenza vaccine is commonly used for clinical prophylaxis in South Africa, we are not aware of previous work to characterise it as an experimental provocation in our population—and this study did not assess the *in vivo* immune response to the influenza vaccine. Similarly, it is unknown whether administering two different annual (2022 and 2023) influenza vaccinations contributed to differences in responses to the influenza vaccination immune challenge, although the statistical analysis did control for this. Follow-up assessments of CPM and TS were assessed at a single time point that aligned with the average peak in IL-6 after the influenza vaccine [53]. However, the cytokine response to influenza vaccine varies between individuals. Consequently, follow up assessments of CPM and TS may have missed the peak inflammatory response to the influenza vaccine challenge in some participants. This study included pain-free adults with varied severity of childhood adversity, on the assumption that these individuals have variable levels of as-yet-unrevealed vulnerability to persistent pain. However, that our sample had no clinical pain may limit the study’s generalisability to clinical pain populations. This sampling decision reflected our priority of understanding how childhood adversity influences vulnerabilities in the nociceptive and inflammatory systems that may lead to persistent pain.

We did not collect self-report data on participants’ ethnicity and ancestry because self-reported ethnicity is a poor proxy for genetic ancestry [43]. Anecdotally, we observed physical characteristics indicating diverse ethnicities and genetic ancestries. The concept of childhood adversity also introduces complexities to the current line of inquiry: adversity is understood differently in different contexts, as shown by the variable performance of the physical neglect subscale of the CTQ-SF, which may reflect poverty rather than neglect [61]. Similarly, corporal punishment is still an accepted disciplinary approach in some South African communities, raising questions about whether all items in the physical abuse subscale reflect physical abuse. The CTQ-SF also has no items for witnessing domestic abuse or witnessing or being a victim of crime, which are common childhood adversities in South Africa. Despite these limitations, the CTQ-SF has good validity [6] and is commonly used in South African research [62]. Hence, it is probably an adequate, albeit imperfect, indicator of childhood adversity in our context.

### Conclusion

The current findings from a heterogenous sample cast doubt on two prominent ideas: that childhood adversity primes the inflammatory system for hyper-responsiveness in adulthood and that nociceptive reactivity is linked to inflammatory reactivity. These important null findings highlight the value of testing research hypotheses in heterogenous samples from diverse contexts to clarify fundamental psychoneuroimmunological mechanisms underlying vulnerability to persistent pain and lay robust foundations of knowledge.

## Data Availability

All data produced in the present study will be made available in a public online repository once the primary manuscript is published.

## Acknowledgements

The authors thank the Clinical Research Centre (Faculty of Health Sciences, University of Cape Town), Henriette Kyepa, and Fathima Docrat for their support with this study, specifically, their role in ordering and storing the influenza vaccines. We thank Srs Rowena Jacobs, Priscilla Nortje, and Dr Rowan Duys for administering the influenza vaccinations and monitoring participants for adverse events. We thank Justin Pead (University of Cape Town) for his technical assistance with automating the high-frequency electrical stimulation trains, Dr Kessie Govender for making the cathodes, and Mathijs Franssen (KU Leuven) for research support.

