## Supplementary files for "Inflammatory reactivity is unrelated to childhood adversity or provoked modulation of nociception"

### 1. CONSORT checklist

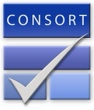

Table S1: CONSORT 2010 checklist of information to include when reporting a randomised trial*

| Section/Topic | Item No | Checklist item | Reported on page No |
| --- | --- | --- | --- |
| Title and abstract | | | |
|  | 1a | Identification as a randomised trial in the title | N/A |
|  | 1b | Structured summary of trial design, methods, results, and conclusions (for specific guidance see CONSORT for abstracts) | Page 4 |
| Introduction | | | |
| Background and objectives | 2a | Scientific background and explanation of rationale | Pages 5 & 6 |
|  | 2b | Specific objectives or hypotheses | Pages 7 & 8 |
| Methods | | | |
| Trial design | 3a | Description of trial design (such as parallel, factorial) including allocation ratio | Page 8 |
|  | 3b | Important changes to methods after trial commencement (such as eligibility criteria), with reasons | Page 8 |
| Participants | 4a | Eligibility criteria for participants | Page 9 |
|  | 4b | Settings and locations where the data were collected | Page 8 |
| Interventions | 5 | The interventions for each group with sufficient details to allow replication, including how and when they were actually administered | Pages 9 – 12 |
| Outcomes | 6a | Completely defined pre-specified primary and secondary outcome measures, including how and when they were assessed | Pages 10 – 12 |
|  | 6b | Any changes to trial outcomes after the trial commenced, with reasons | Page 8 |
| Sample size | 7a | How sample size was determined | Page 15 |
|  | 7b | When applicable, explanation of any interim analyses and stopping guidelines | N/A |
| Randomisation: |  |  |  |
| Sequence generation | 8a | Method used to generate the random allocation sequence | N/A |
|  | 8b | Type of randomisation; details of any restriction (such as blocking and block size) | N/A |
| Allocation concealment mechanism | 9 | Mechanism used to implement the random allocation sequence (such as sequentially numbered containers), describing any steps taken to conceal the sequence until interventions were assigned | N/A |
| Implementation | 10 | Who generated the random allocation sequence, who enrolled participants, and who assigned participants to interventions | N/A |
| Blinding | 11a | If done, who was blinded after assignment to interventions (for example, participants, care providers, those assessing outcomes) and how | Page 14 |
|  | 11b | If relevant, description of the similarity of interventions | N/A |
| Statistical methods | 12a | Statistical methods used to compare groups for primary and secondary outcomes | Pages 15 & 16 |
|  | 12b | Methods for additional analyses, such as subgroup analyses and adjusted analyses | Pages 15 & 16 |
| Results | | | |
| Participant flow (a diagram is strongly recommended) | 13a | For each group, the numbers of participants who were randomly assigned, received intended treatment, and were analysed for the primary outcome | Page 16 |
|  | 13b | For each group, losses and exclusions after randomisation, together with reasons | Page 16 |
| Recruitment | 14a | Dates defining the periods of recruitment and follow-up | Page 8 |
|  | 14b | Why the trial ended or was stopped | N/A |
| Baseline data | 15 | A table showing baseline demographic and clinical characteristics for each group | Page 17 |
| Numbers analysed | 16 | For each group, number of participants (denominator) included in each analysis and whether the analysis was by original assigned groups | Page 17 |
| Outcomes and estimation | 17a | For each primary and secondary outcome, results for each group, and the estimated effect size and its precision (such as 95% confidence interval) | Pages 18 – 20 |
|  | 17b | For binary outcomes, presentation of both absolute and relative effect sizes is recommended | N/A |
| Ancillary analyses | 18 | Results of any other analyses performed, including subgroup analyses and adjusted analyses, distinguishing pre-specified from exploratory | Page 21 |
| Harms | 19 | All important harms or unintended effects in each group (for specific guidance see CONSORT for harms) | Supplementary file: Table S5 |
| Discussion | | | |
| Limitations | 20 | Trial limitations, addressing sources of potential bias, imprecision, and, if relevant, multiplicity of analyses | Page 26 |
| Generalisability | 21 | Generalisability (external validity, applicability) of the trial findings | Pages 25 & 26 |
| Interpretation | 22 | Interpretation consistent with results, balancing benefits and harms, and considering other relevant evidence | Pages 23 & 24 |
| Other information | | |  |
| Registration | 23 | Registration number and name of trial registry | Page 8 |
| Protocol | 24 | Where the full trial protocol can be accessed, if available | Page 8 |
| Funding | 25 | Sources of funding and other support (such as supply of drugs), role of funders | Page 2 |

Citation: Schulz KF, Altman DG, Moher D, for the CONSORT Group. CONSORT 2010 Statement: updated guidelines for reporting parallel group randomised trials. BMC Medicine. 2010;8:18.
© 2010 Schulz et al. This is an Open Access article distributed under the terms of the Creative Commons Attribution License (<http://creativecommons.org/licenses/by/2.0>), which permits unrestricted use, distribution, and reproduction in any medium, provided the original work is properly cited.

*We strongly recommend reading this statement in conjunction with the CONSORT 2010 Explanation and Elaboration for important clarifications on all the items. If relevant, we also recommend reading CONSORT extensions for cluster randomised trials, non-inferiority and equivalence trials, non-pharmacological treatments, herbal interventions, and pragmatic trials. Additional extensions are forthcoming: for those and for up-to-date references relevant to this checklist, see [www.consort-statement.org](http://www.consort-statement.org).

### 2. Protocol deviations

Table S2: Summary of protocol deviations and justifications for the deviations.

| **Protocol deviation number** | **Stated in the protocol** | **Deviation from the protocol** | **Justification(s) for deviation** |
| --- | --- | --- | --- |
| 1 | We will determine the area under the curve formed by the three estimates of surface area over time since HFS, as the dependent variable for hypothesis 3. The within-participant peak of surface area of secondary hypersensitivity provides insight into the peak severity of secondary hypersensitivity; therefore, we will also compute and report this at the group level. | We included all three measures of surface area across the three timepoints for each participant in our statistical analysis. | We consulted a statistician who informed us that using area under the curve in this study has 3 limitations: (i) it reduces the intra-individual variability in the data, (ii) it reduces the power because of the removing the repeated measures, and (iii) area under the curve is not a stable measurement for just three repeated measures. |
| 2 | We calculate the combined arithmetic mean for the three sets of the two pinprick stimuli to obtain a stable estimate of baseline sensory ratings to mechanical punctate stimulation.  The magnitude of secondary hypersensitivity will be calculated for each post-HFS time point by subtracting ratings before HFS from each rating after HFS. Again, we will determine the area under the curve formed by the three estimates of magnitude of secondary hypersensitivity over time since HFS, as the dependent variable for hypothesis 3. The within-participant peak magnitude of secondary hypersensitivity provides insight into the peak severity of secondary hypersensitivity; therefore, we will also compute and report this at the group level. | We included ratings for each stimuli, at baseline and all three follow-up points for each participant in our statistical analysis. | We consulted a statistician who informed us that using area under the curve in this study has 3 limitations: (i) it reduces the intra-individual variability in the data, (ii) it reduces the power because of the removing the repeated measures, and (iii) area under the curve is not a stable measurement for just three repeated measures. |
| 3 | Broken blinding will be assessed using James’ Blinding Index. | Broken blinding will be assessed using chi-squared test. |  |
| 4 | PRK (data analyst) will remain blinded to natures of the 3 groups to undertake blinded model specification, assessment, and interpretation of the best fitting models. To achieve this, VJM will use a script that is stored outside the analysis repository, out of access to PRK, before formal statistical analyses, and PRK will have no access to preliminary data analysis files or discussion of the preliminary analysis. PRK will choose, assess, and confirm the final statistical models and provide conclusions about each study hypothesis, and this document will be locked online, before PRK is unblinded for further discussion and more detailed interpretation of the study findings. | During data analysis, the data analyst was unblinded participants’ CA group allocations. | It was deemed unnecessary for the data analyst to be blinded to group allocation because CA group allocation was merely used to ensure recruitment of a cohort representing a range of CA history. CA group status was not included in any of the statistical analyses. |

### 3. Estimates of cytokine levels

To estimate the levels of IL-1b and IL-6, we undertook the following calculations to obtain a standard curve and estimate the observed values for each panel in a reproducible manner. First, the background fluorescence was subtracted from the fluorescence observed to obtain the net florescence of each well. Second, we used the standards data to obtain an appropriate standard curve. To obtain an initial standard curve, we fitted a quadratic model to each standard, using expected concentrations (dependent variable) and net observed fluorescence (independent variable). We assessed the accuracy of this model by plotting observed vs expected standards data against a line showing model-predicted values, on both a linear scale and a log-log scale. Poor fit was apparent at the lower end of the range. We assessed this further by calculating the model-predicted values for each standard value, and the %CV between the model-predicted values and observed values. The %CV increased as the values dropped, showing poor model fit towards the lower end of the range.

To address this systematic bias in model fit, we defined a weight for each analyte and each standard as the inverse of the expected concentration (e.g. weightIL6 = 1/(expected-valueIL6)) and then fitted a quadratic model as before, but now with weighting that increased the influence of each data point in inverse proportion to its expected value. We assessed the accuracy of this weighted quadratic model by plotting observed vs expected data against a line showing model-predicted values, on both a linear scale and a log-log scale. The fit was better at the lower end of the range, although the %CV between the model-predicted values and observed values still increased (but less dramatically) as the values dropped. The weighted quadratic model was carried forward as the standard curve.

Third, we used our weighted quadratic model (standard curve) to predict the true values of the test samples from the observed net fluorescence values. Estimates that fell outside the bounds of the expected range for the panel were flagged as “out of range” and are reported for each analyte (see Results). Given that the fitted model suggested linearity of the data, we interpolated new values for each samples flagged as ‘out of range’ as follows:

- For values above the expected range, we used the net fluorescence value at the maximum of the expected range (i.e. net fluorescence for Standard 1) plus half the difference between the highest (S1) and second-highest (S2) standards.
- For values below the expected range, we used the net fluorescence values at the minimum of the expected range (i.e. predicted value for Standard 6) minus half the difference between the lowest (S6) and second-lowest (S5) standards.
- We then used the weighted quadratic model to predict the values at each of these net fluorescence values. Finally, the mean of the two predicted values was calculated and then multiplied by 30, to account for the sample dilutions.

### 4. Method for determining individual detection threshold for the high-frequency electrical stimulation

The individual detection threshold was determined using an established calibration process in which single electrical stimuli (pulse width 2 ms) were delivered with an adaptive staircase method, starting from zero and increasing in 0.1 mA increments (steps) until the participant reported feeling the electrical stimulus (usually felt as a “tiny prick”) at the cathode site. Next, the intensity was decreased by steps of half the previous step size (i.e. 0.05 mA) until the participant reported no longer feeling the electrical stimulus. Next, the intensity was increased by steps of half the previous step size (i.e. 0.025 mA) until the participant reported feeling the electrical stimulus again. This final reporting of feeling the electrical stimulus was used as the individual detection threshold.

### 5. Outcome measures for potential confounders

#### 5.1 Positive childhood experiences

Participants reported on positive childhood experiences using the Positive Childhood Experiences Questionnaire [57]. This questionnaire has seven items to which participants endorsed answers using a 5-point Likert scale (0 – never; 1 – rarely; 2 – sometimes; 3 – often; 4 – very often). The total score was computed as the sum of all seven item scores. Positive childhood experiences may buffer the effects of adverse childhood experiences [57]. We tested whether score on the positive childhood experiences questionnaire influenced the relationship childhood adversity and inflammatory reactivity.

#### 5.2 Long-term stress

Participants completed the 10-item Perceived Stress Scale [58] by rating each item (e.g. *In the last month, how often have you been able to control the irritations in your life)* on a 5-point Likert scale (1 – never; 2 – almost never; 3 – sometimes; 4 – fairly often; 5 – very often). The total score was computed as the sum of all 10 item scores. Long-term stress alters inflammatory reactivity, as seen by high levels of IL-6 in people exposed to chronic stress [59-65]. Further, individuals exposed to long-term stress of caring for a spouse with dementia showed lower levels of IL-2 after influenza vaccine immune provocation than age- and sex-matched controls [66]. Therefore, we tested whether long-term stress was a potential confounder of inflammatory reactivity, surface area and magnitude of secondary hypersensitivity, CPM, and/or TS.

#### 5.3 Depression and anxiety

We screened for a lifetime history of diagnosed major depressive disorder, as well as screened for current symptoms of depression and anxiety using the well-validated and reliable Patient Health Questionnaire-4 [67]. This questionnaire has 4-items to which answers were endorsed using a 4-point Likert scale (0 – not at all; 1 – several days; 2 – more than half the days; 3 – nearly every day). The sum of the first two items screen for anxiety and the sum of the last two items screen for depression (>3 is considered positive for either of these subscales). Depression alters inflammatory reactivity, as seen by a meta-analysis of data from 3212 people reporting higher levels of IL-6 and TNF-α, as well as other cytokines, in peripheral blood of people with major depressive disorder, than in healthy controls [68]. Anxiety is also associated with higher levels of IL-6 and TNF-α [69]. Therefore, we tested whether depression and/or anxiety (total screening score on Patient Health Questionnaire-4, and previous diagnosis of major depressive disorder) were potential confounders of inflammatory reactivity, surface area and magnitude of secondary hypersensitivity, CPM, and/or TS.

#### 5.4 Asthma

Participants self-reported history of diagnosed asthma. Asthma alters inflammatory reactivity, as seen by preliminary, unpublished data from our research team. Further, asthma history is associated with reduced levels of the largely anti-inflammatory cytokine IL-10 [70] in LPS-stimulated blood culture, and asthma severity is negatively associated with levels of IL-10 [71]. Therefore, we tested whether diagnosed asthma was a potential confounder of inflammatory reactivity.

#### 5.5 COVID-19 infection

Participants self-reported history of COVID-19 infection in the six months preceding participation. In participants with a history of COVID-19 infection, we collected data on the timing and severity of known COVID-19 infection and severity of long-COVID symptoms. Recent COVID-19 infection may alter inflammatory reactivity, and could confound interpretation of IL-6 and TNF-α levels in stimulated and unstimulated blood samples. COVID-19 infection stimulates a cytokine-driven inflammatory response that can build into a ‘cytokine storm’ with profound and lasting effects [72, 73]. Further, persistent symptoms after acute COVID-19 infection, termed “long-COVID” are associated with pro-inflammatory cytokine activation [74, 75]. Therefore, history of COVID-19 infection may be associated with inflammatory reactivity. Unfortunately, we were unable to account for undiagnosed COVID-19 infection, which is suspected to be common although the inflammatory consequences are unknown. We tested whether COVID-19 infection was a potential confounder of inflammatory reactivity.

#### 5.6 Chronic and recent acute illnesses

Participants self-reported chronic illnesses and any recent acute infections in the six months preceding participation, and we tested whether chronic illnesses (e.g. HIV) and recent acute illnesses (e.g. influenza) was a potential confounder of inflammatory reactivity.

#### 5.7 Sleep

Participants reported sleep quality using the Pittsburgh Sleep Quality Index [76], which consists of 19 self-rated items, the sum of which provides a total score out of 21. Sleep deprivation and/or disturbance can alter inflammatory and neural reactivity, as shown by data from humans under resting and challenged states. A large meta-analysis of data from 3000 participants reported a strong positive association between recent sleep disturbance and circulating IL-6 levels [77]. Additionally, poor sleep quality was associated with higher levels of IL-6 after *in vivo* stimulation using LPS in African American women [78]. Further, one night of sleep deprivation increased the area of experimentally induced secondary hyperalgesia in healthy male, but not female, subjects [79]. Therefore, we tested whether sleep deprivation (total score out of 21) was a potential confounder of inflammatory reactivity, surface area and magnitude of secondary hypersensitivity, CPM, and/or TS.

### 6. Sensitivity power analysis

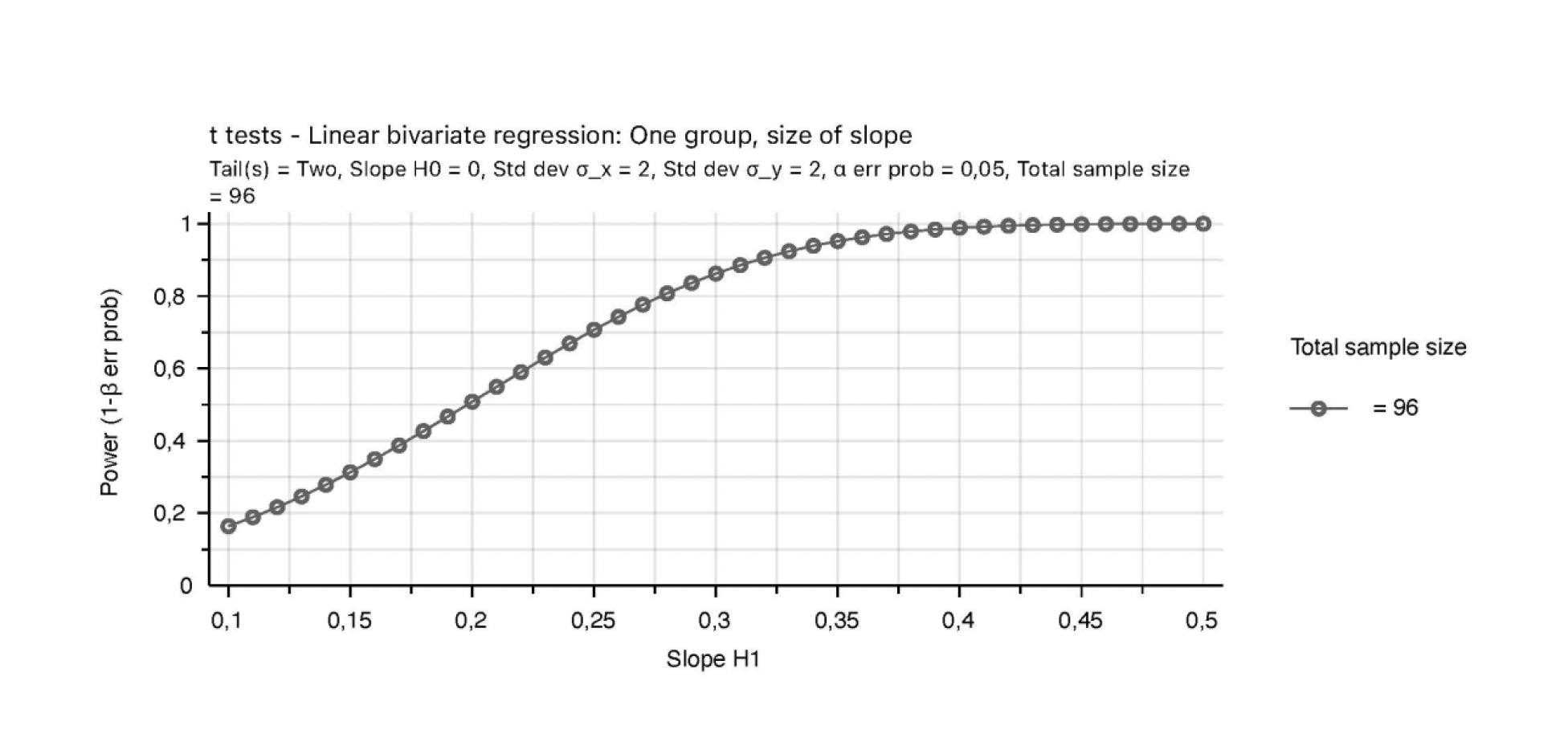

Figure S1: **Sensitivity power analysis** to estimated that our final sample size (n=96) provided a priori power to detect an effect size of 0.275 with power 0.8 and alpha 0.05.

### 7. Descriptive statistics

Table S3: Descriptive statistics

| **Characteristics** | **Full sample**  **N = 96** | **History of mild childhood adversity (CTQ-SF score: 25 – 36)**  **N = 32** | **History of moderate childhood adversity (CTQ-SF score: 37 – 67)**  **N = 32** | **History of severe childhood adversity (CTQ-SF score: 68 – 125)**  **N = 32** | **p-value for between-group differences** |
| --- | --- | --- | --- | --- | --- |
| **Positive childhood experiences** (Positive Childhood Experiences Questionnaire)^‡^ | 15.9 (±6.6) | 22.1 (±5.2) | 18.6 (±3.8) | 11.3 (±4.8) | **<0.001** |
| **Long-term stress** (Perceived Stress Scale)^§^ | 31.0 (28.0 – 33.0) | 30.0 (28.8 – 32.5) | 30.0 (28.0 – 33.0) | 32 (29.8 – 35.3) | 0.32 |
| **Depression and anxiety** | | | | | |
| Diagnosis of major depressive disorder | 7 (7.29%) | 2 (6.3%) | 1 (3.1%) | 4 (12.4%) | - |
| Patient Health Questionnaire-4^¶^ | 4 .0 (1.0 – 6.0) | 1.5 (0.0 – 5.0) | 2.0 (1.0 – 4.0) | 5.0 (3.0 – 8.0) | **0.01** |
| Depression subscale^#^ | 2.0 (0.0 – 3.0) | 0.0 (0.0 – 2.3) | 1.0 (0.0 – 2.0) | 2.0 (1.0 – 4.0) | **0.02** |
| Anxiety subscale^#^ | 2.0 (1.0 – 3.3) | 1.5 (0.0 – 2.0) | 2.0 (1.0 – 2.0) | 3.0 (2.0 – 4.0) | **0.04** |
| **Diagnosis of asthma** | 4 (4.2%) | 1 (3.1%) | 2 (6.3%) | 1 (3.1%) | - |
| **COVID-19 infection in preceding 6 months** | 7 (7.29%) | 3 (9.4%) | 4 (12.4%) | 0 | - |
| **Chronic and recent acute illness** |  | | | | |
| Diagnosed chronic illness(es) | 11 (11.46%)  HIV n = 5  Hypertension n = 2  Polycystic ovarian syndrome n = 2  Chronic allergic rhinitis n = 1  Inverse psoriasis n = 1 | | | | |
| Recent acute illness(es) in preceding 6 months | 56 (58.3%) | 20 (62.5%) | 18 (56.2%) | 18 (56.2%) | 0.84 |
| **Sleep** (Pittsburgh Sleep Quality Index)** | 7.0 (5.0 – 10.0) | 5.0 (3.8 – 9.3) | 6.5 (5.0 – 10.0) | 8.5 (5.8 – 11.3) | 0.095 |
| **Self-reported side effects of influenza vaccine** | | | | | |
| Participants reporting any side effects | 48 (50.0%) | 19 (59.4%) | 15 (46.9%) | 14 (43.8%) | 0.42 |
| List of side effects | Pain at the vaccination site n = 39 (40.6%)  Fatigue n = 12 (12.5%)  Headache n = 11 (1.5%)  Flu-like symptoms n = 5 (5.2%)  Body aches n = 6 (6.3%)  Pain in axilla ipsilateral to vaccine site n = 2 (2.1%)  Swollen axillary lymph nodes ipsilateral to vaccine site n = 2 (2.1%)  Chest pain n = 1  Night sweats n = 1  Dry mouth n = 1  Moody n = 1  Fever n = 1  Stomach ache n = 1  Muscle twitches n = 1 | Pain at the vaccination site n = 17 (53.1%)  Fatigue n = 2 (6.3%)  Headache n = 4 (12.5%)  Flu-like symptoms n = 2 (6.3%)  Body aches n = 1 (3.1%)  Swollen axillary lymph nodes ipsilateral to vaccine site n = 2 (6.3%)  Chest pain n = 1 | Pain at the vaccination site n = 12 (37.5%)  Fatigue n = 6 (18.8%)  Headache n = 4 (12.5%)  Flu-like symptoms n = 1 (3.1%)  Body aches n = 3 (9.4%)  Night sweats n = 1  Dry mouth n = 1 | Pain at the vaccination site n = 10 (31.2%)  Fatigue n = 4 (12.5%)  Headache n = 3 (9.4%)  Flu-like symptoms n = 2 (6.3%)  Body aches n = 2 (6.2%)  Pain in axilla ipsilateral to vaccine site n = 2 (6.2%)  Moody n = 1  Fever n = 1  Stomach ache n = 1  Muscle twitches n = 1 |  |
| Overall rated severity of side effects^††^ | 1.9 (±1.1) | 1.6 (±0.9) | 2.0 (±1.1) | 2.3 (±1.1) | 0.21 |

^‡^ 49 (of 96) participants completed the Positive Childhood Experiences Questionnaire; possible total score range: 0 – 28

^§^ Possible total score range for the Perceived Stress Scale: 10 – 50

^¶^ 48 (of 96) participants completed the Patient Health Questionnaire-4; possible total score range: 0 – 12

^#^ Possible total score range for the depression and anxiety subscales of the PHQ-4: 0 – 6

** Possible total score range for the Pittsburgh Sleep Quality Index: 0 – 21

^††^ Possible score range for rated severity of side effects: 5-point Likert scale, where 1 = “very mild and 5 = “very severe”)

Data are presented as median (IQR), mean (±SD), or n (%)

### 8. Manipulation checks

Table S4: Summary of the main effect of condition (before vs during cold water immersion) on pressure pain threshold (PPT) at each test site. PPT = pressure pain threshold.

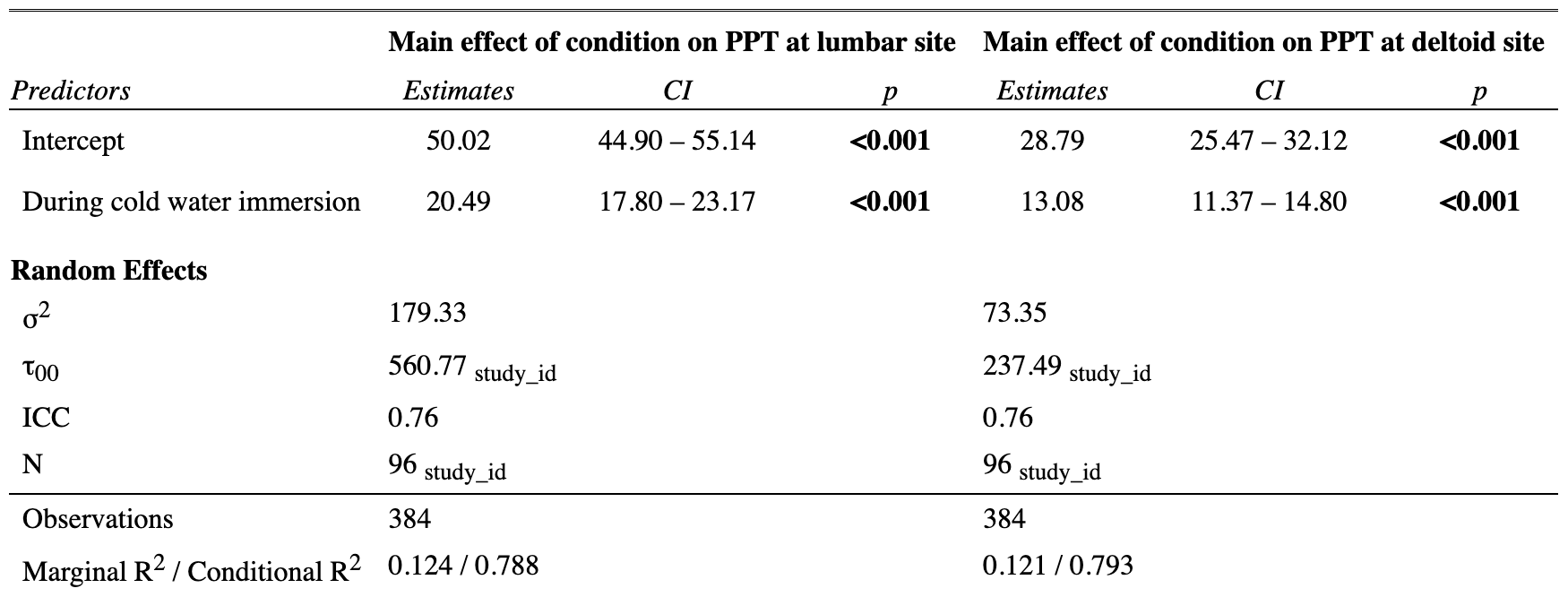

Table S5: Summary of main effect of condition (1^st^ vs 16^th^ stimulation) on SPARS ratings to mechanical stimuli.

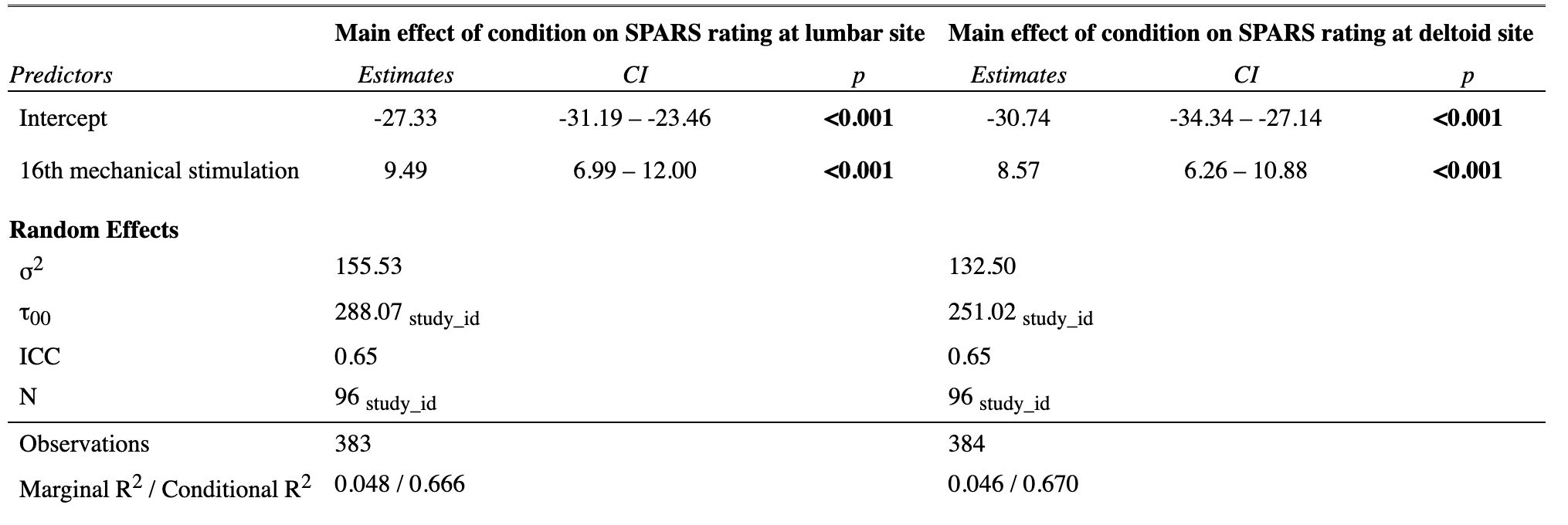

### 9. Hypothesis 1: relationship between childhood adversity and provoked cytokine expression

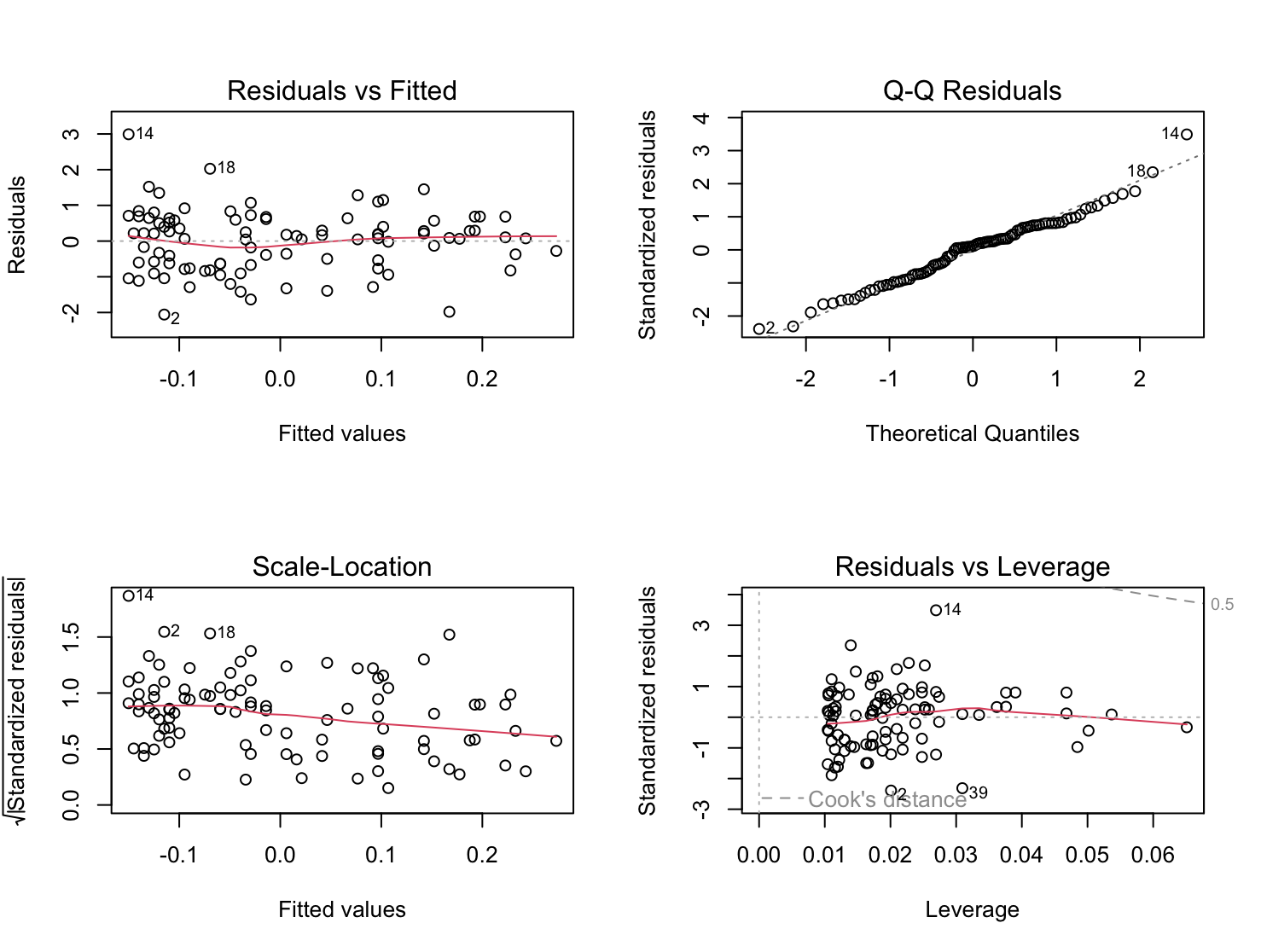

Figure S2: Plots of linear regression model assumptions for hypothesis 1.

Table S6: Summary of unadjusted and adjusted models for hypothesis 1. CTQ-SF = Childhood Trauma Questionnaire-short form; PSQI = Pittsburgh Sleep Quality Index; PSS = Perceived Stress Scale.

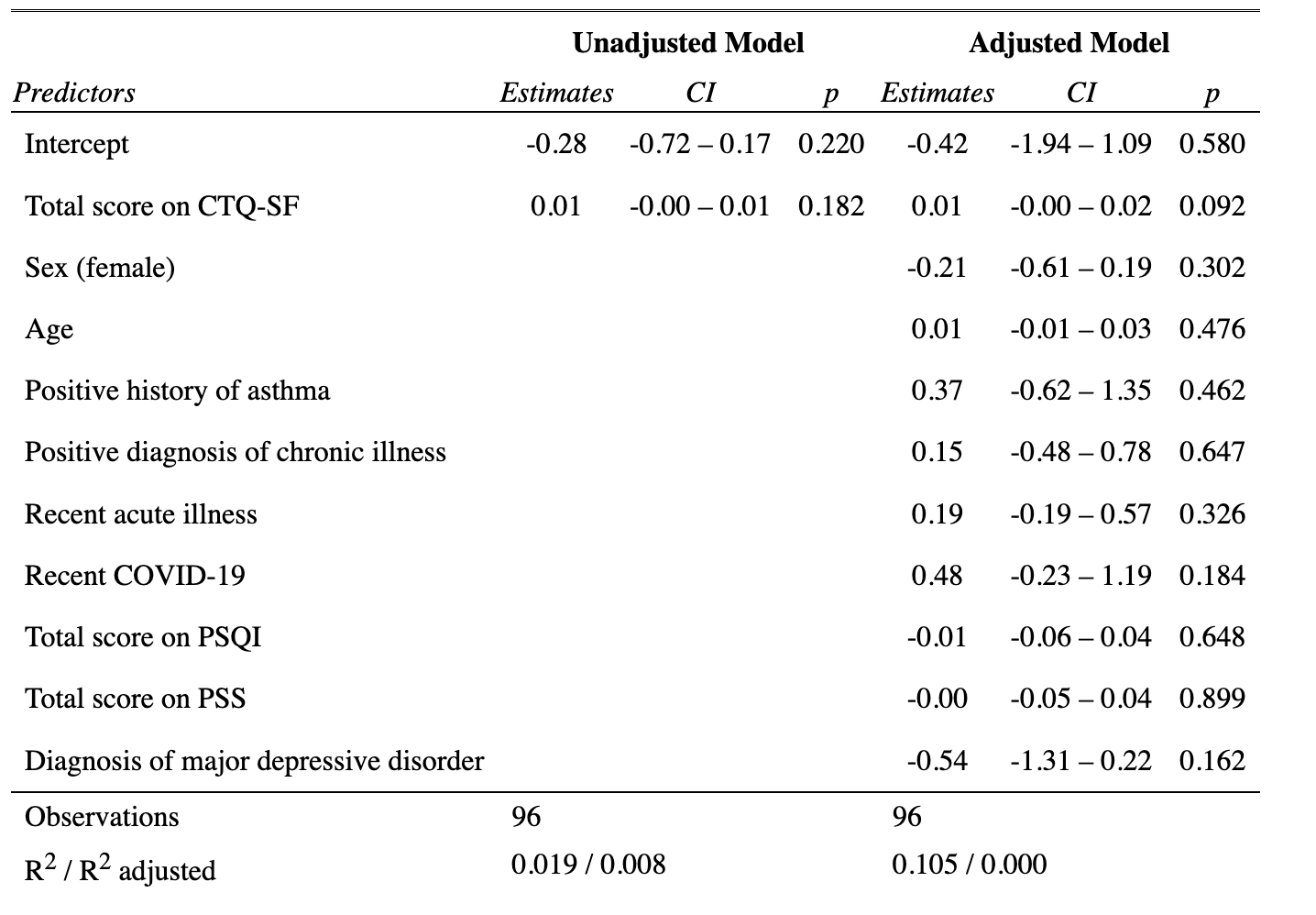

### 10. Hypothesis 2: relationship between provoked cytokine expression and induced secondary hyperalgesia

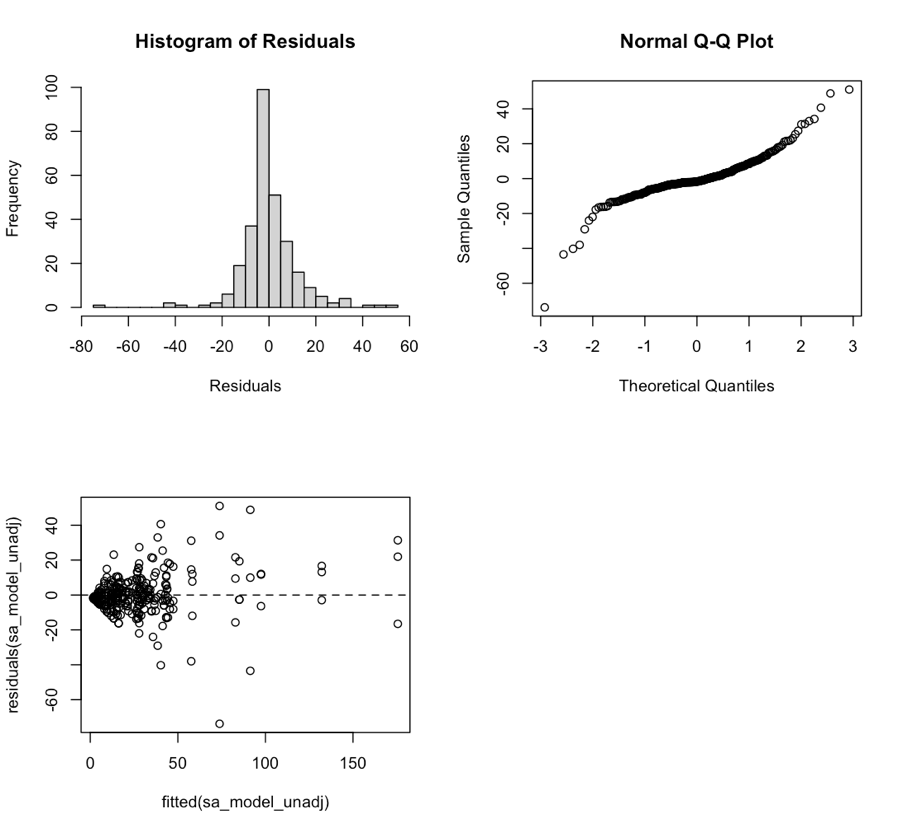

Figure S3: Model assumptions for conventional linear regression model for the surface area of secondary hyperalgesia.

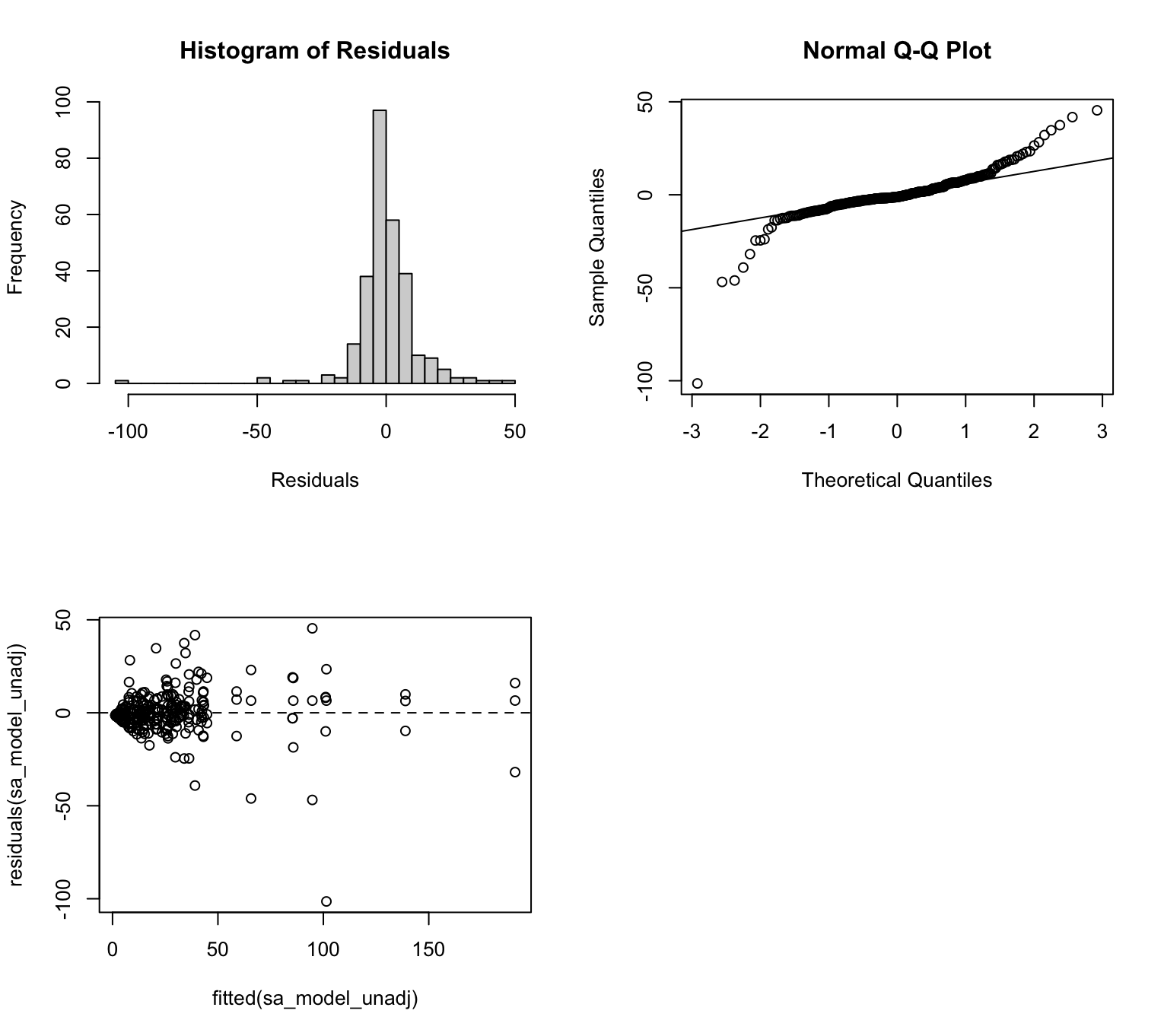

Figure S4: Model assumptions for robust linear regression model for the surface area of secondary hyperalgesia.

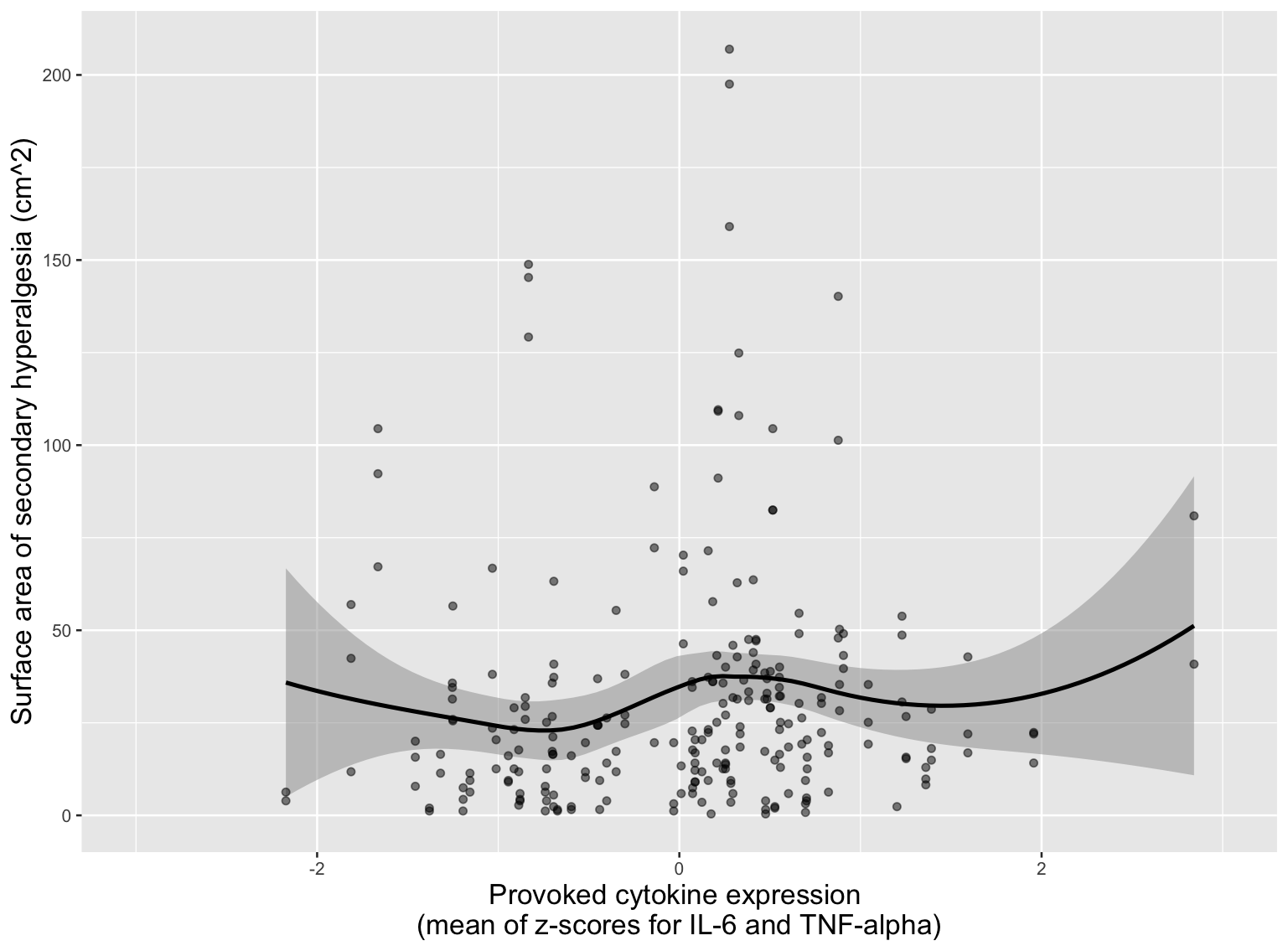

Figure S5: Relationship between provoked cytokine expression and surface area, where the area is > 0 cm^2^, of secondary hyperalgesia after neural provocation (i.e. HFS induction).

Table S7: Summaries of unadjusted and adjusted hurdle models for hypothesis 2. Outcome: surface area of secondary hyperalgesia. PSS = Perceived Stress Scale; HFS = high-frequency electrical stimulation; PSQI = Pittsburgh Sleep Quality Index; MDD = major depressive disorder.
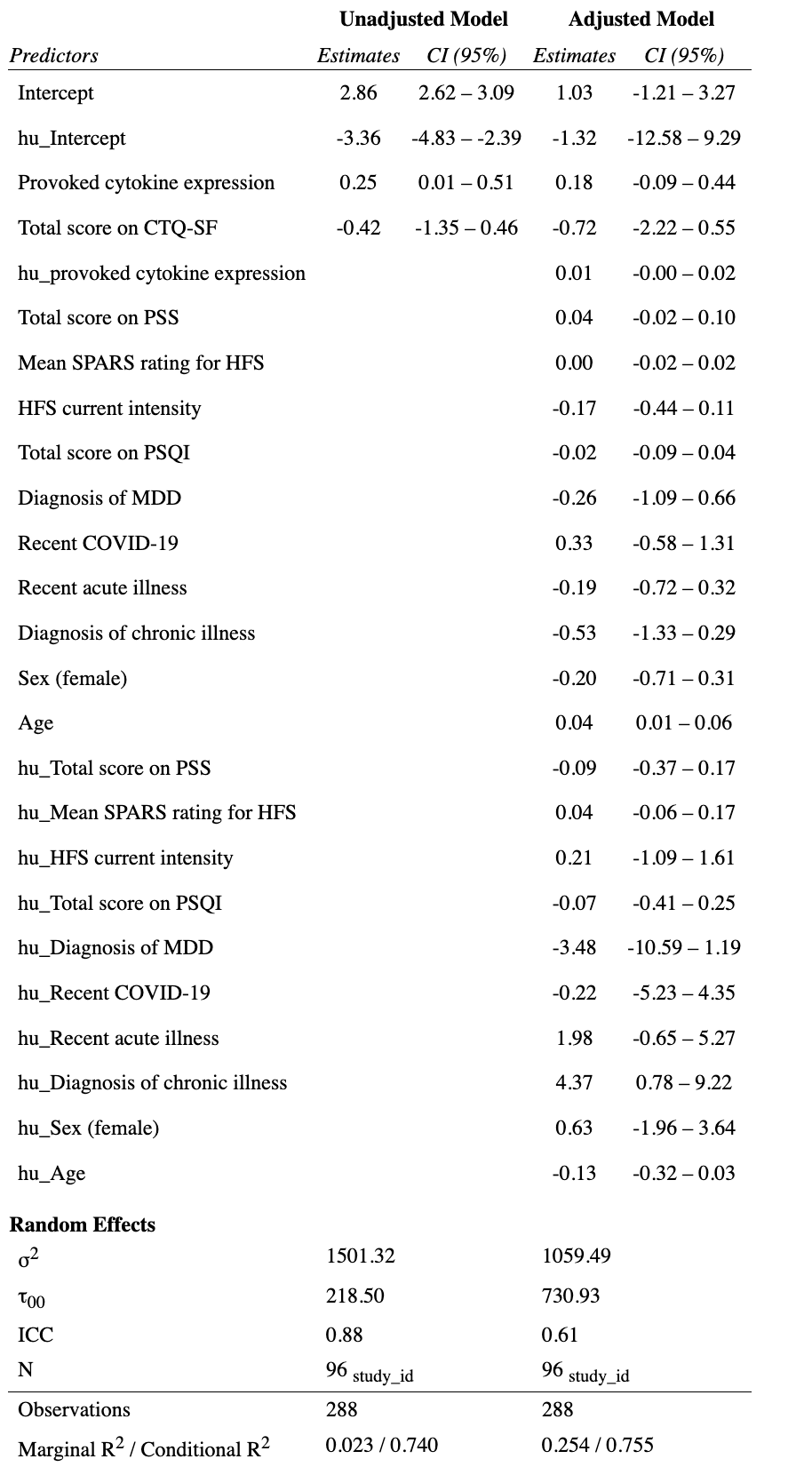

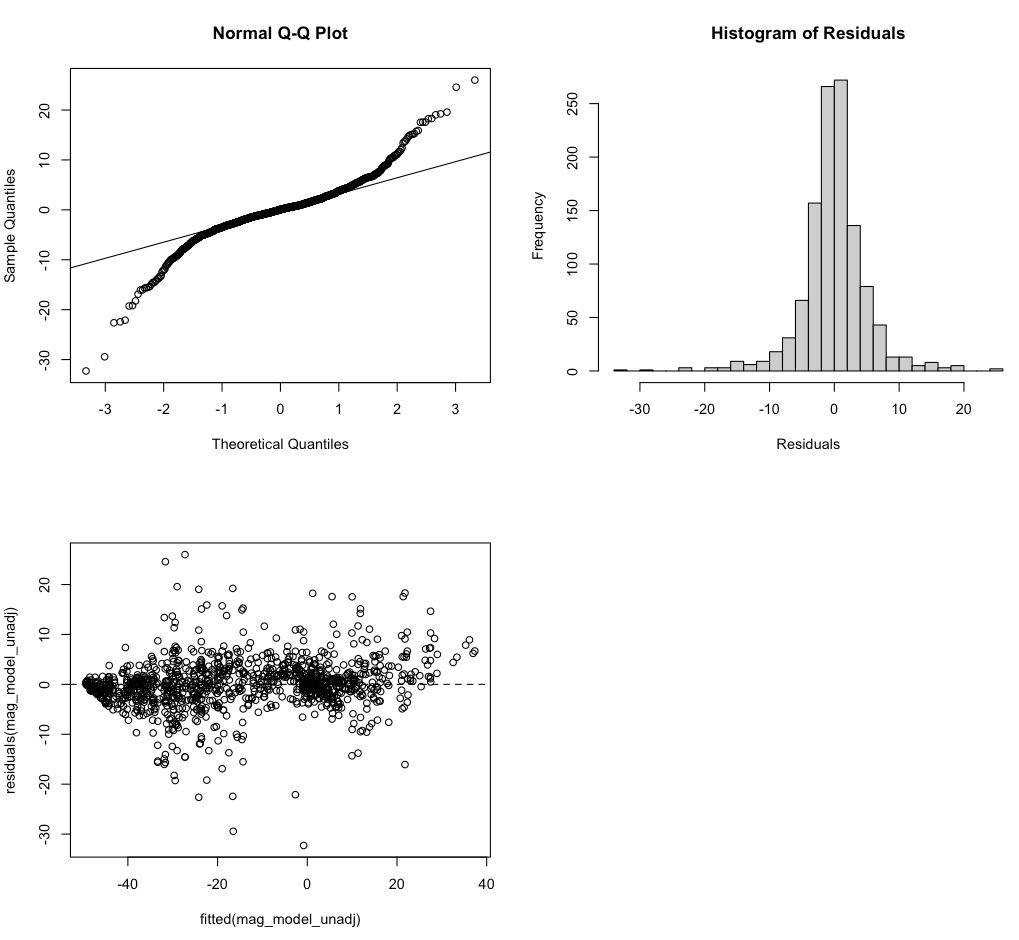

Figure S6: Model assumptions for conventional linear regression model for magnitude of secondary hyperalgesia

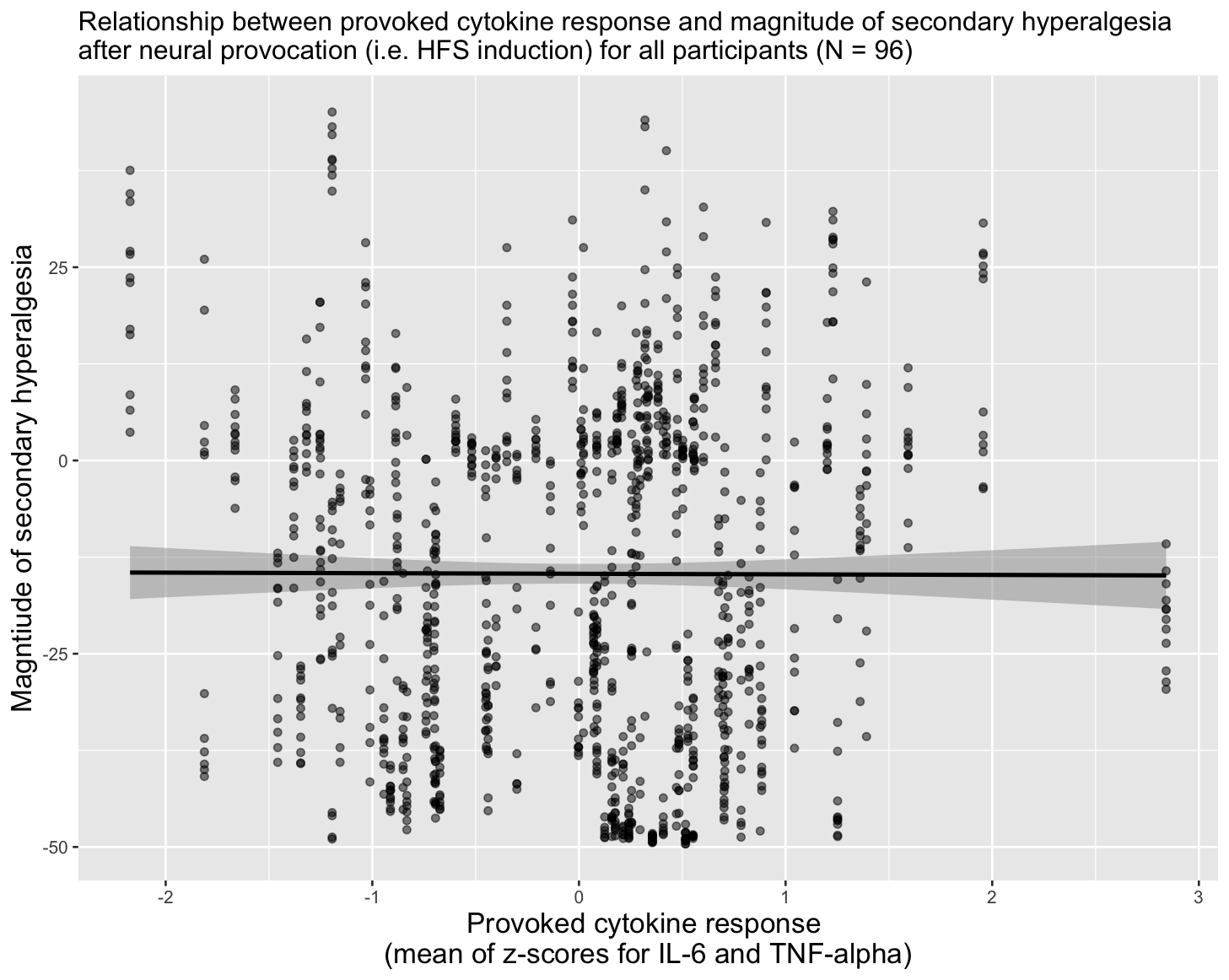

Figure S7: Relationship between provoked cytokine expression and magnitude of secondary hyperalgesia after neural provocation (i.e. HFS induction) for all participants (n=96).

Table S8: Summary of unadjusted and adjusted models for hypothesis 2. Outcome: magnitude of secondary hyperalgesia. HFS = high-frequency electrical stimulation; CTQ-SF = Childhood Trauma Questionnaire-Short Form; PSS = Perceived Stress Scale; PSQI = Pittsburgh Sleep Quality Index.

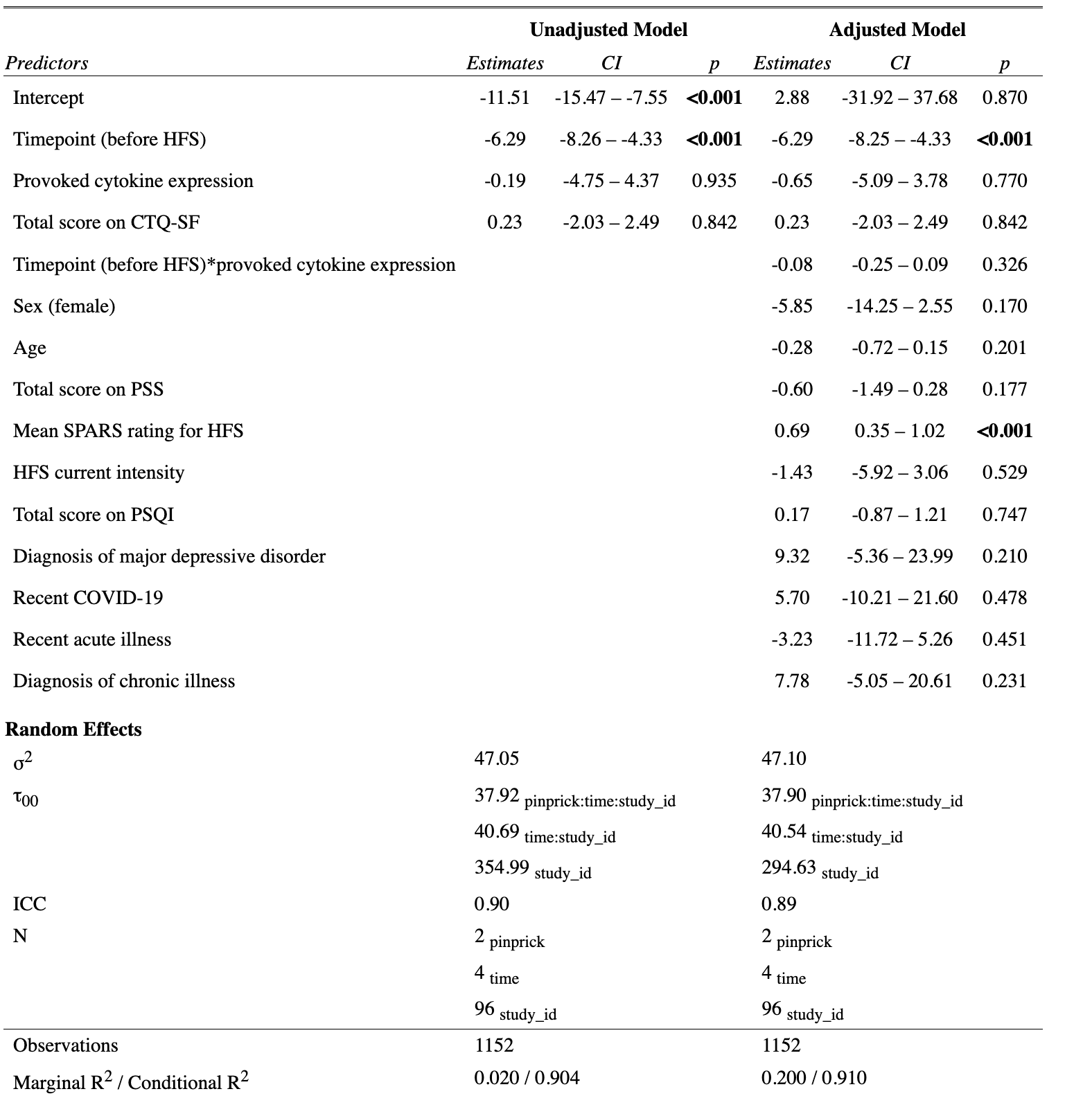

### 11. Hypothesis 3: relationship between provoked cytokine expression and change in CPM and TS

Table S9: Summary of the main effect of session (before vs after influenza vaccination) on conditioned pain modulation (CPM) at each test site.

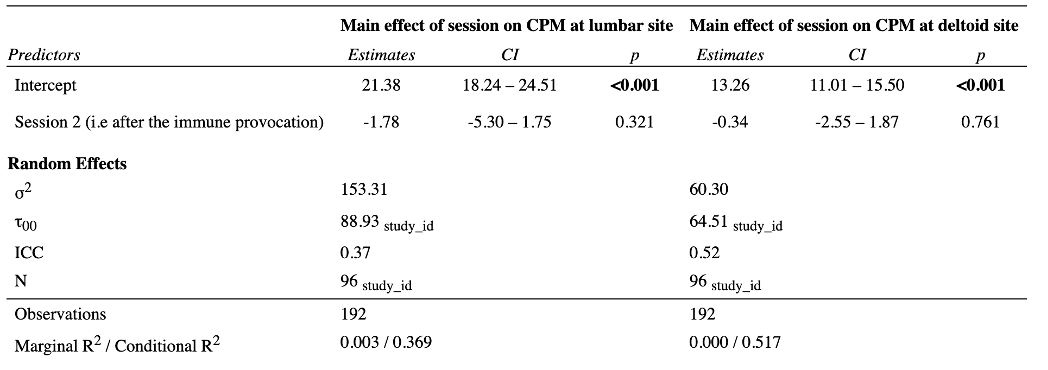

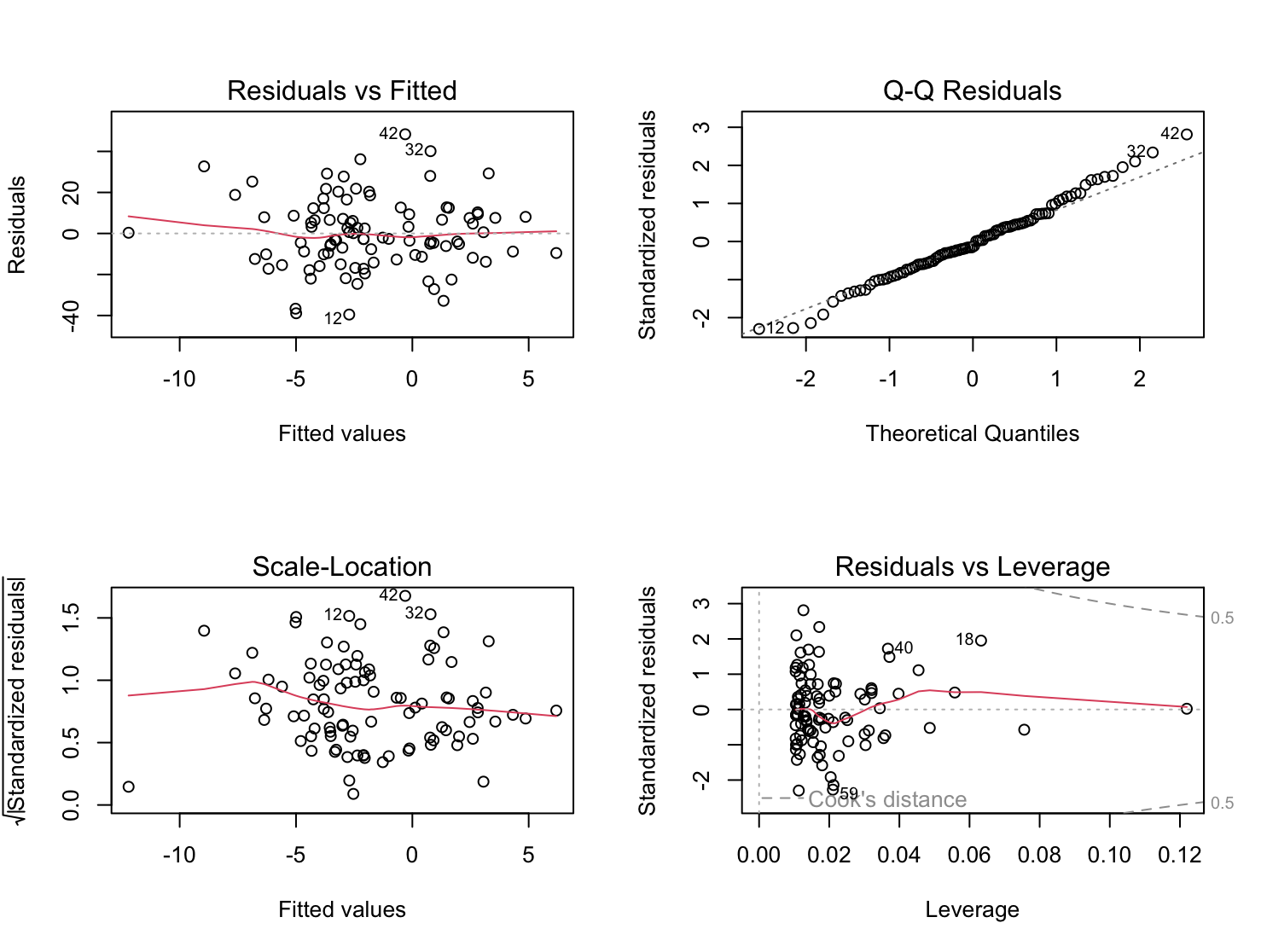

Figure S8: Plots of linear regression model assumptions for hypothesis 2. Outcome: change in CPM; test site: lumbar.

Table S10: Summary of unadjusted and adjusted models for hypothesis 3. Outcome: change in conditioned pain modulation; test site: lumbar. CTQ-SF = Childhood Trauma Questionnaire-Short Form; PSS = Perceived Stress Scale; PSQI = Pittsburgh Sleep Quality Index.

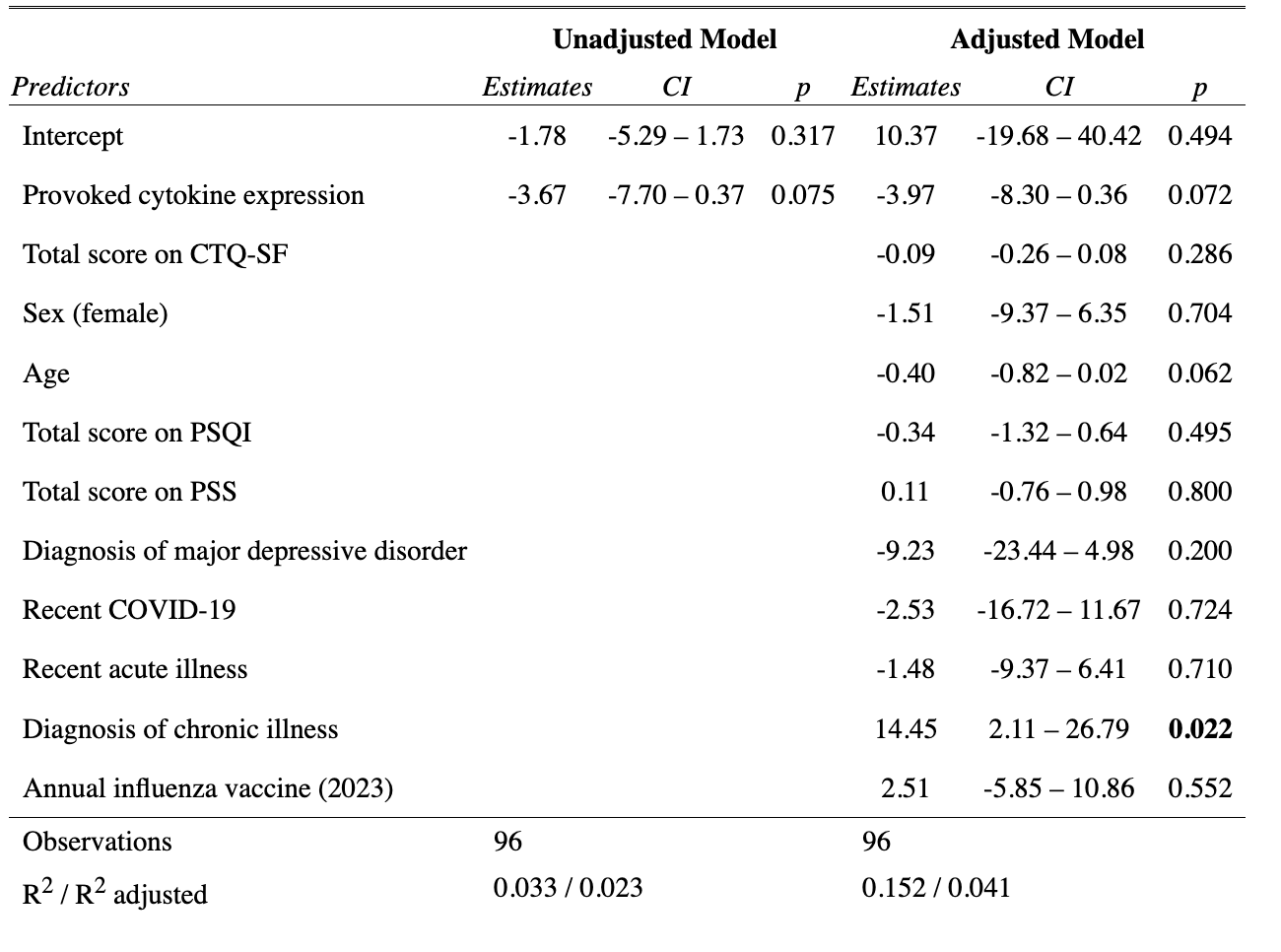

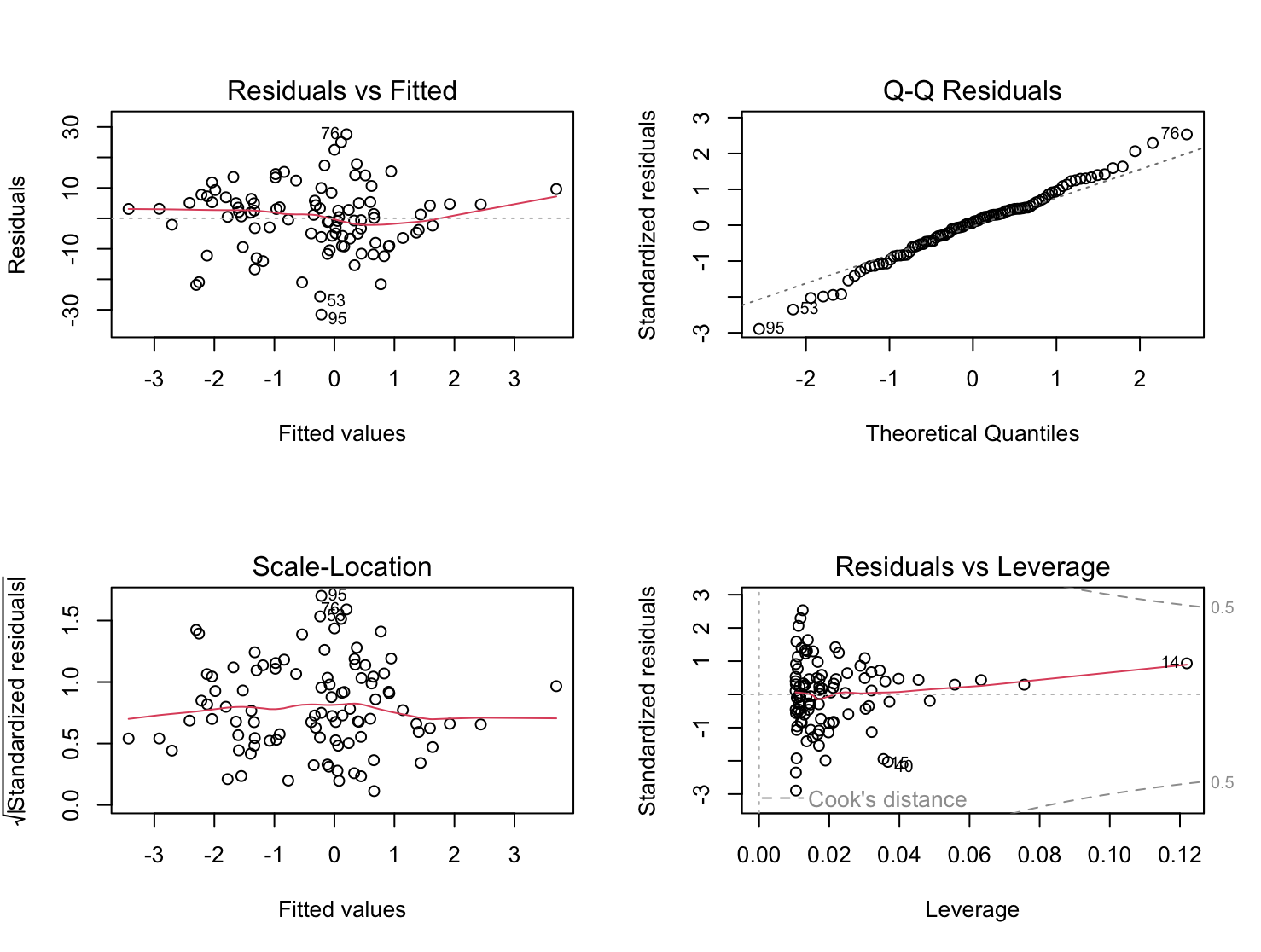

Figure S9: Plots of linear regression model assumptions for hypothesis 3. Outcome: change in CPM; test site: deltoid.

Table S11: Summary of unadjusted and adjusted models for hypothesis 3. Outcome: change in conditioned pain modulation; test site: deltoid. CTQ-SF = Childhood Trauma Questionnaire-Short Form; PSS = Perceived Stress Scale; PSQI = Pittsburgh Sleep Quality Index.

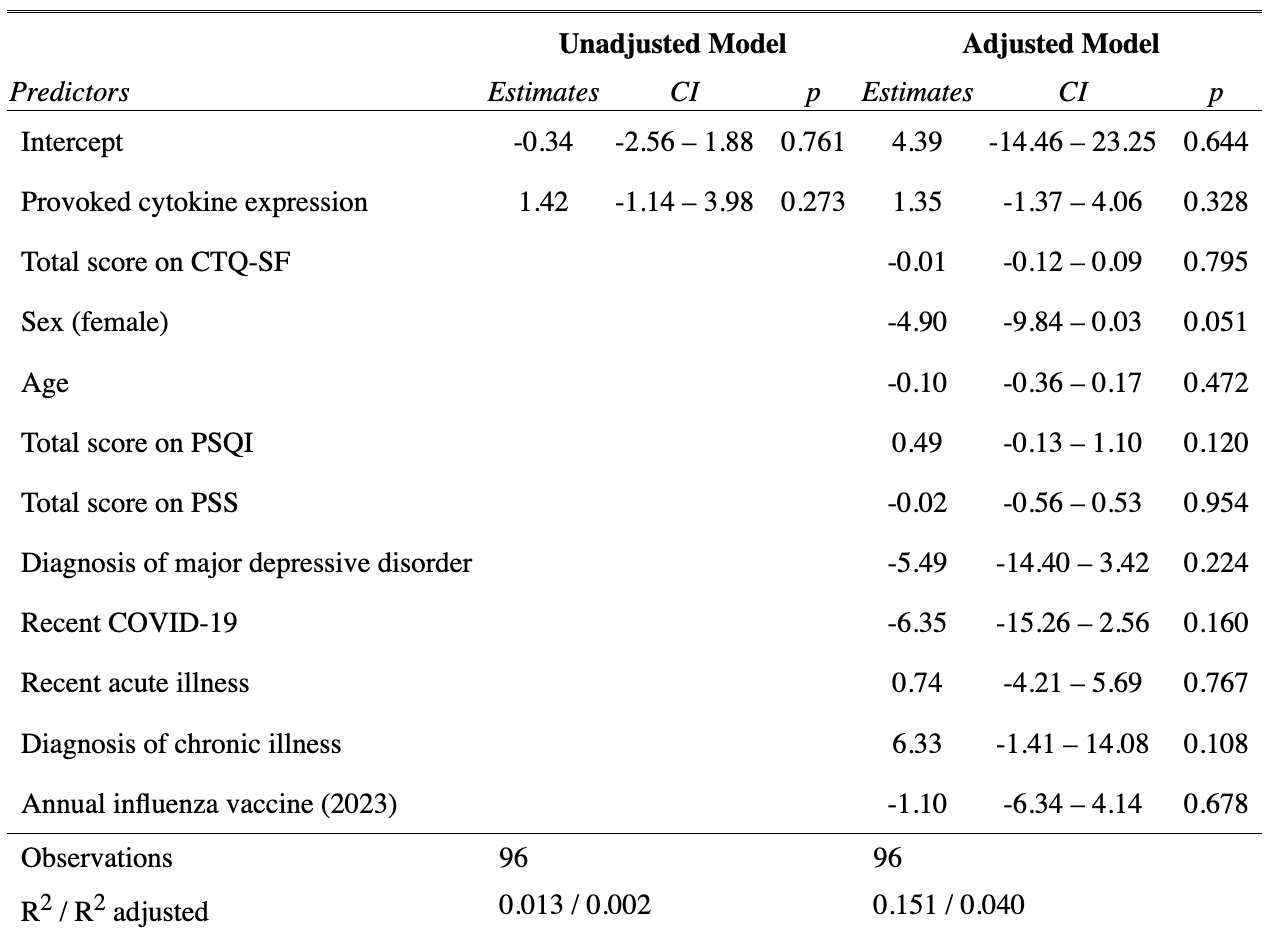

#### 11.1 Relationship between provoked cytokine expression and change in CPM stratified by sex

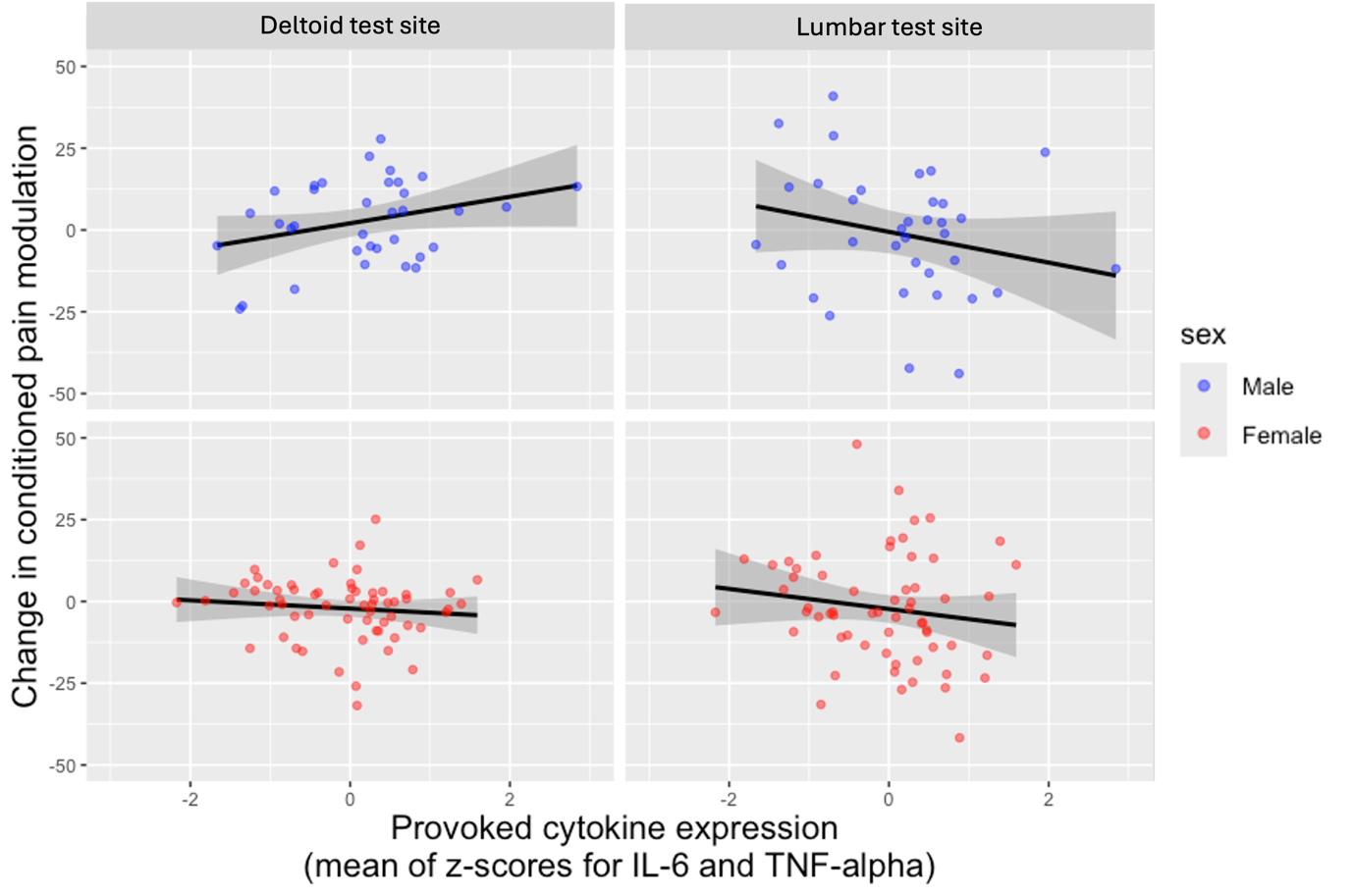

Figure S10: The relationship between provoked cytokine expression and change in conditioned pain modulation stratified by sex at the deltoid and lumbar test sites.

Table S12: Summary of the main effect of session (before vs after influenza vaccination) on temporal summation (TS) at each test site.

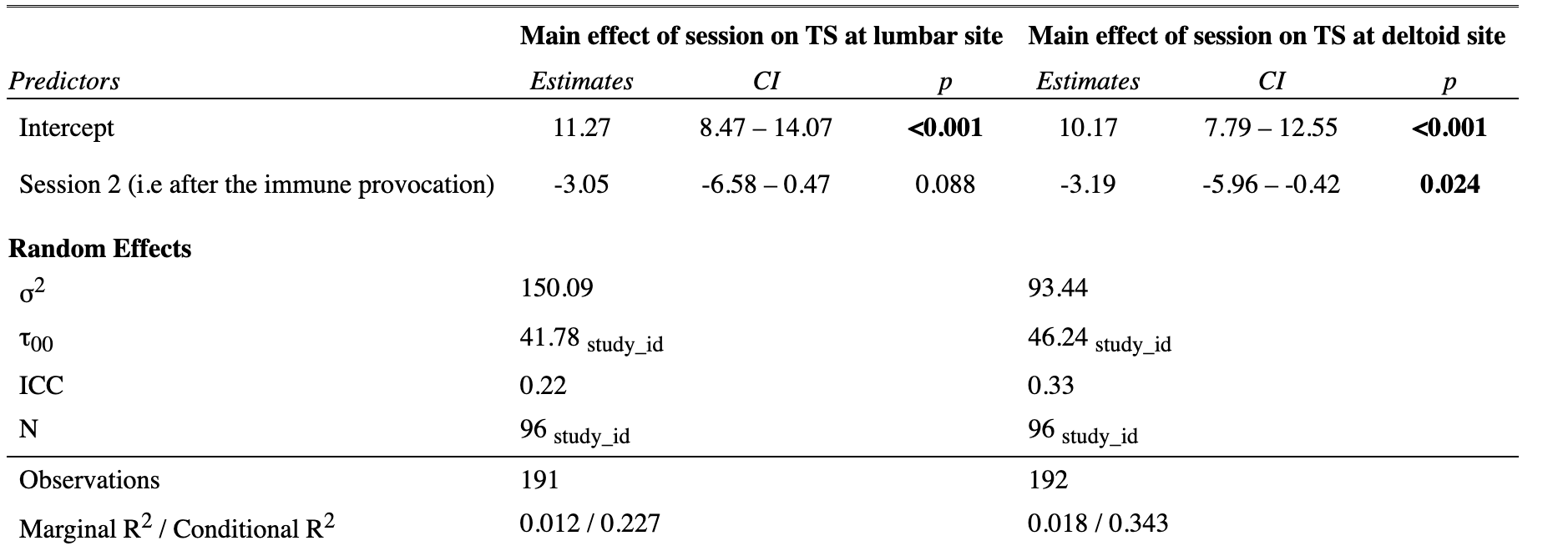

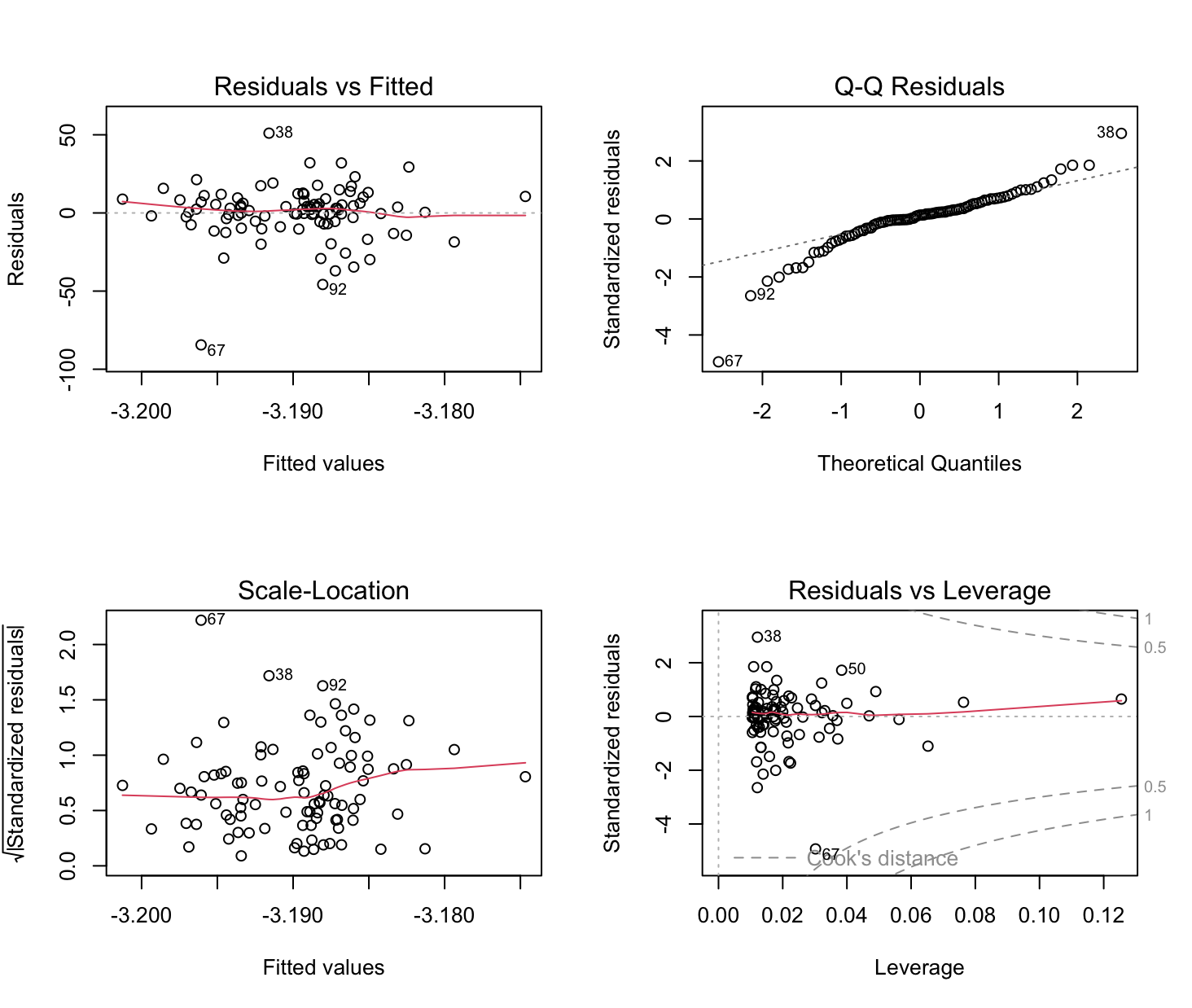

Figure S11: Plots of linear regression model assumptions for hypothesis 3. Outcome: change in TS; test site: lumbar

Table S13: Summary of unadjusted and adjusted models for hypothesis 3. Outcome: change in temporal summation; test site: lumbar. CTQ-SF = Childhood Trauma Questionnaire-Short Form; PSS = Perceived Stress Scale; PSQI = Pittsburgh Sleep Quality Index.

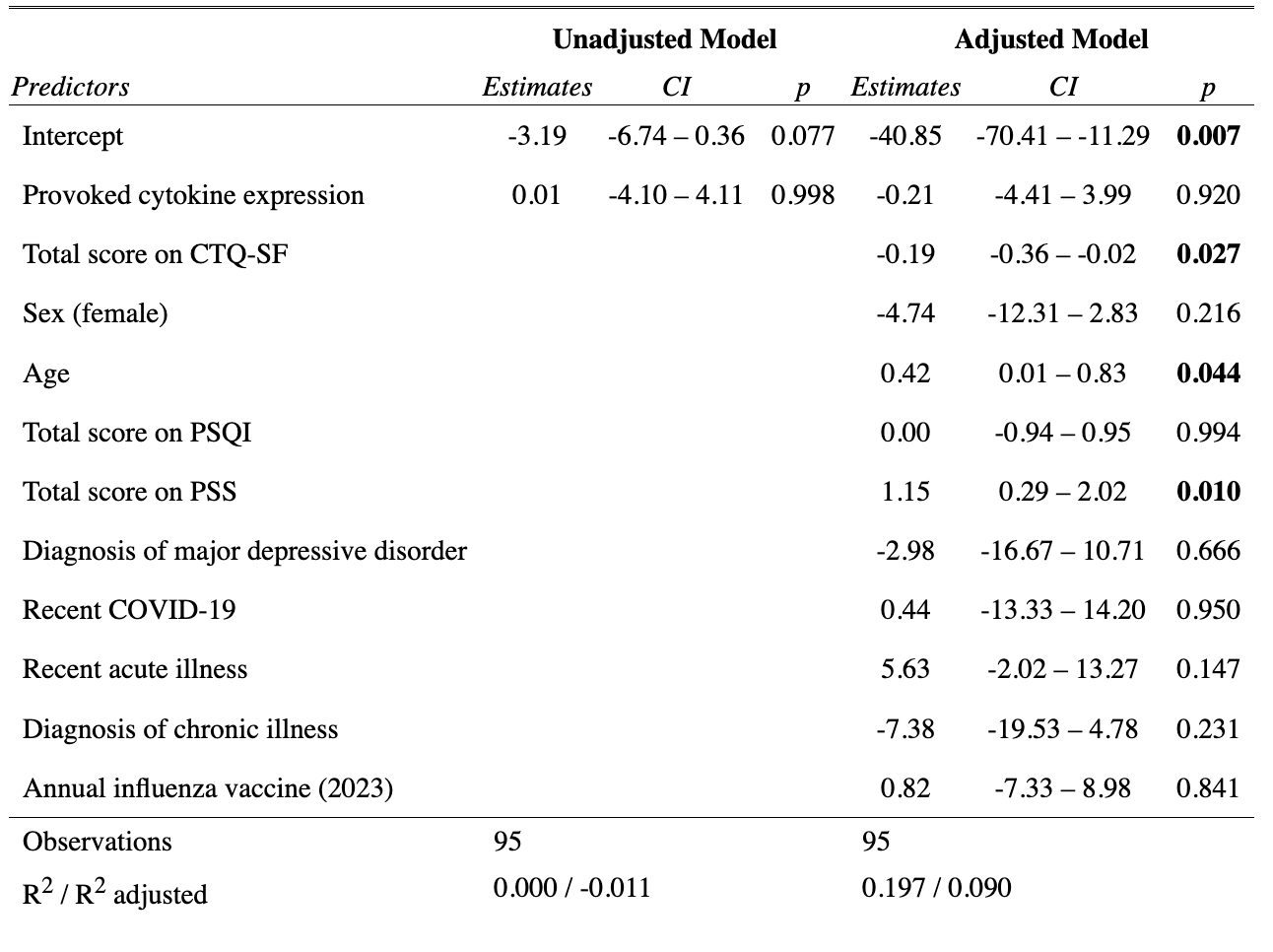

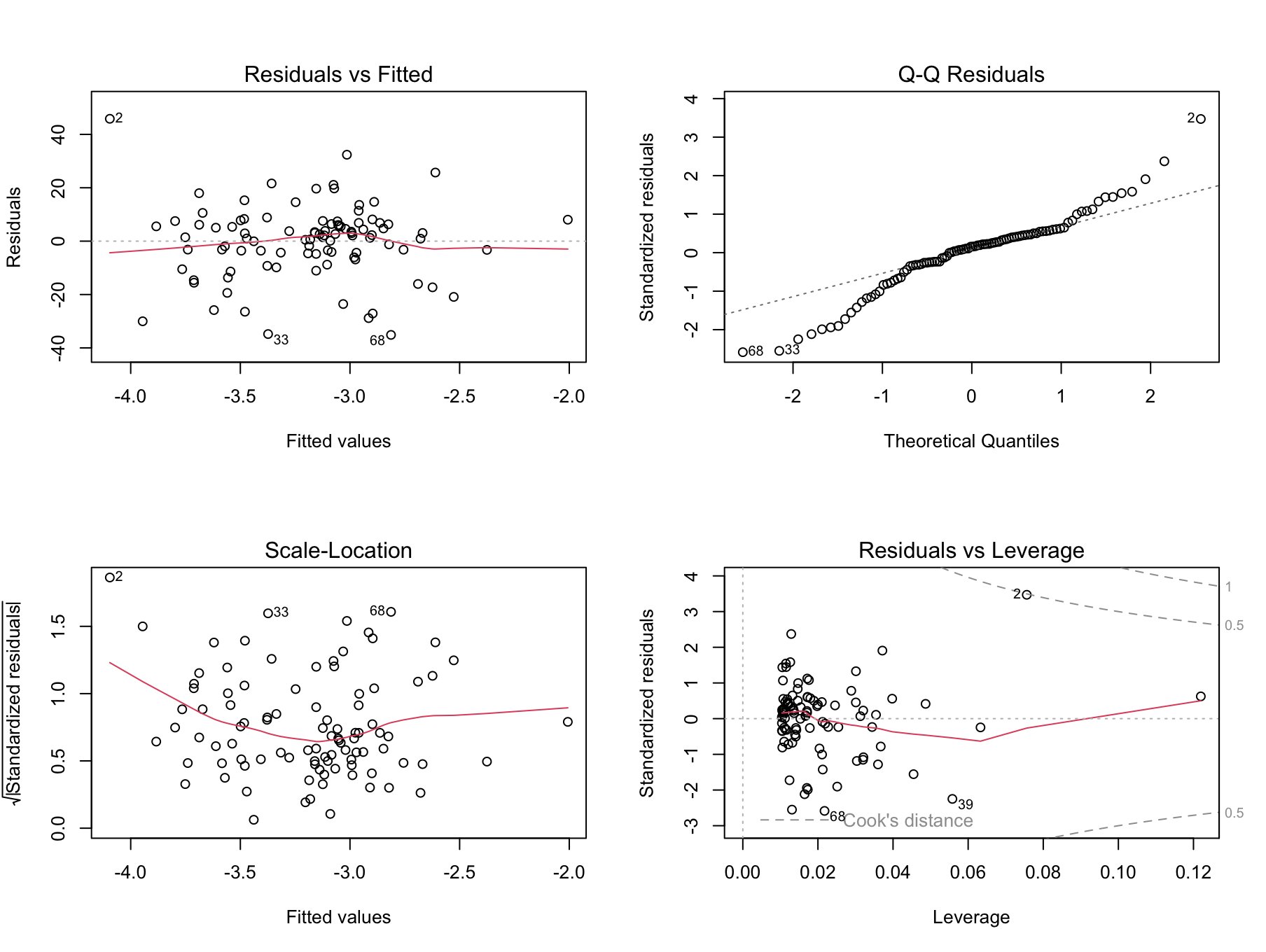

Figure S12: Plots of linear regression model assumptions for hypothesis 3. Outcome: change in TS; test site: deltoid.

Table S14: Summary of unadjusted and adjusted models for hypothesis 2. Outcome: change in temporal summation; test site: deltoid. CTQ-SF = Childhood Trauma Questionnaire-Short Form; PSS = Perceived Stress Scale; PSQI = Pittsburgh Sleep Quality Index.

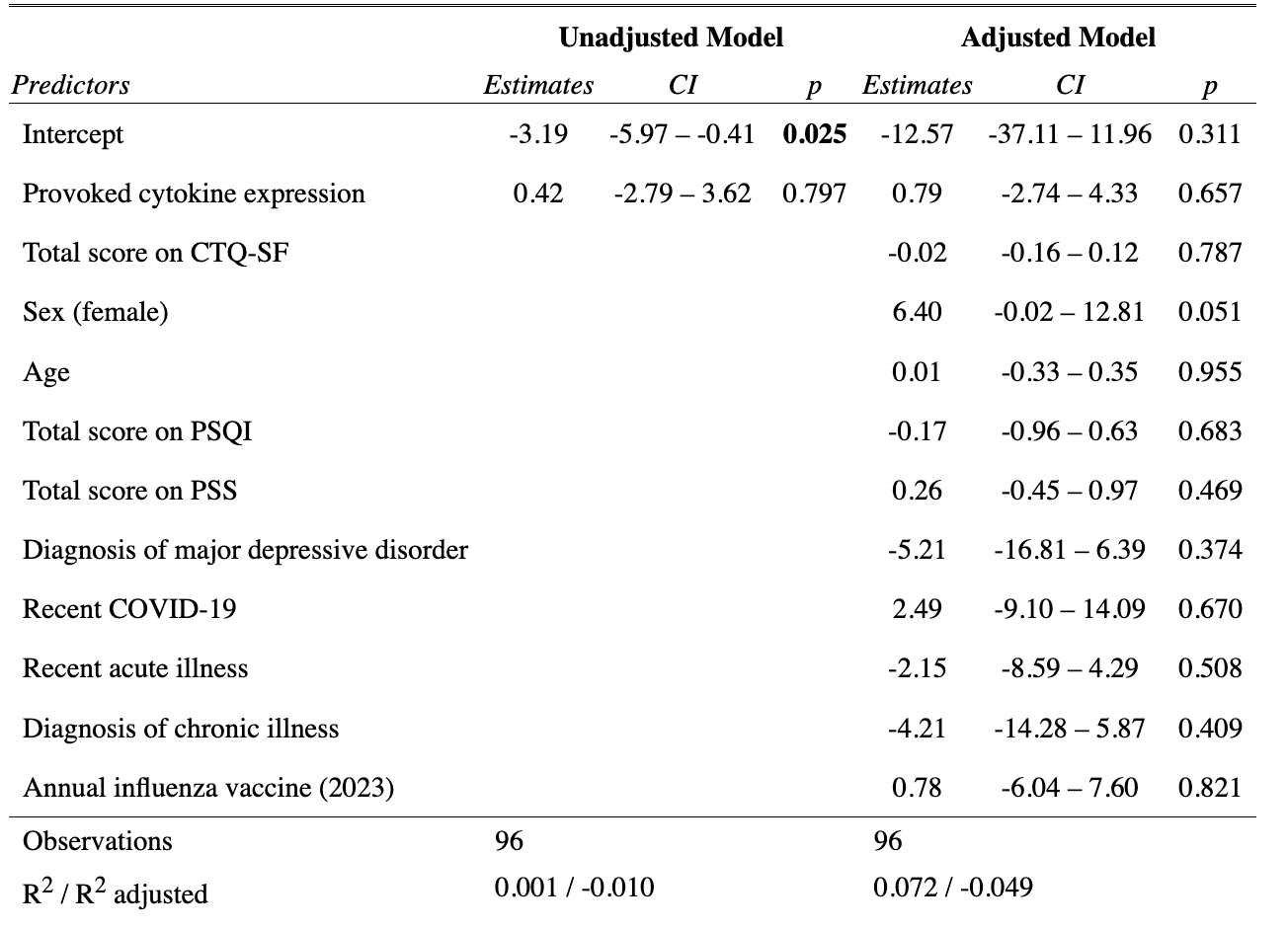

### 12. Blinding assessment

Table S15: Summary of unadjusted and covariate-adjusted models for hypothesis 1 for the full sample and sensitivity analyses excluding 2 (of 96) participants who were unblinded to hypothesis 1. PSS = Perceived Stress scale; PSQI = Pittsburgh Sleep Quality Index.

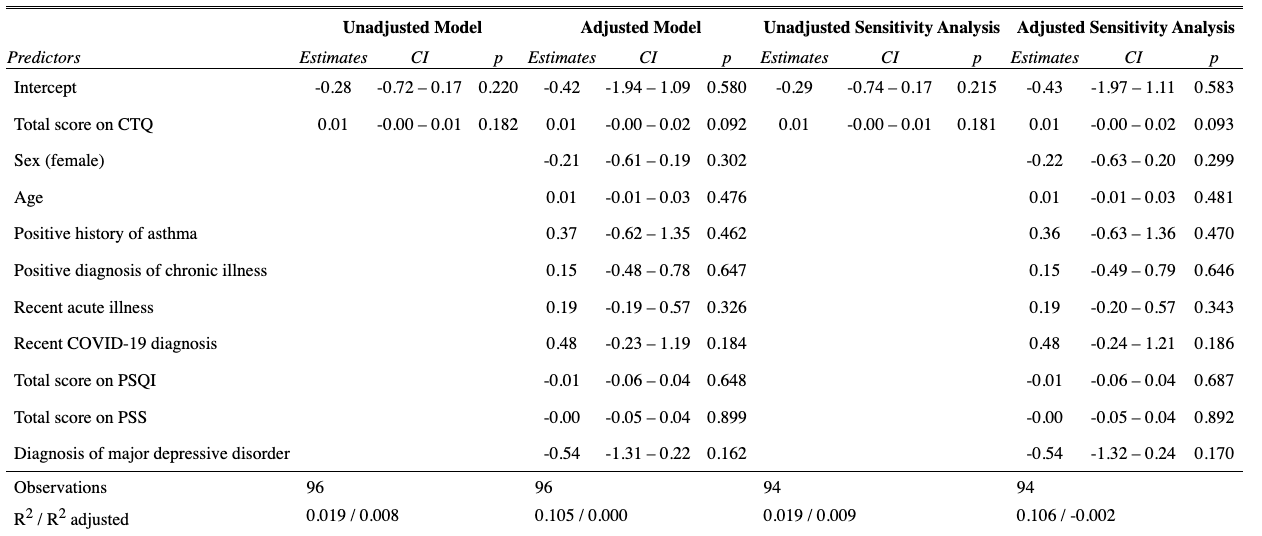

Table S16: Summary of unadjusted and covariate-adjusted models for hypothesis 3 (change in CPM at the lumbar site) for the full sample and sensitivity analyses excluding 4 (of 96) participants who were unblinded to hypothesis 3. PSS = Perceived Stress scale; PSQI = Pittsburgh Sleep Quality Index.

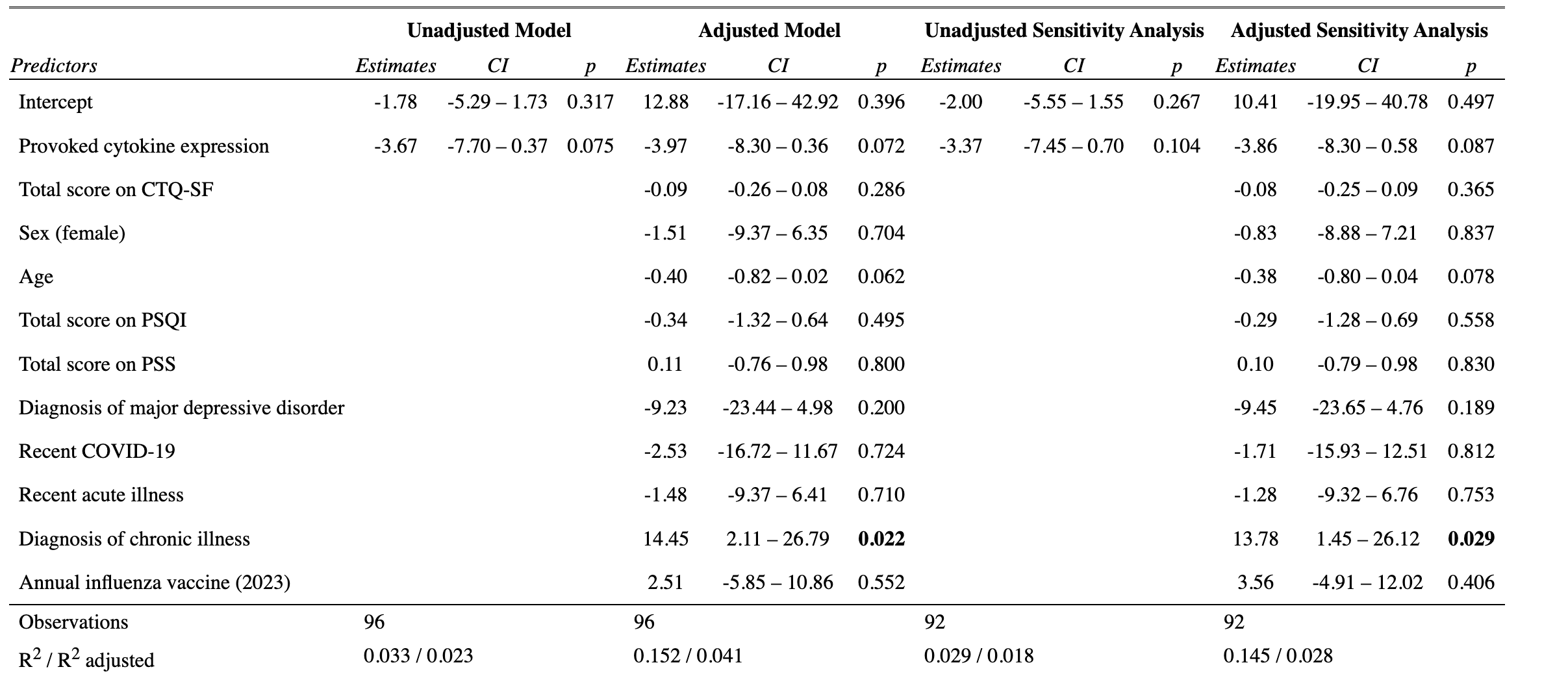

Table S17: Summary of unadjusted and covariate-adjusted models for hypothesis 3 (change in CPM at the deltoid site) for the full sample and sensitivity analyses excluding 4 (of 96) participants who were unblinded to hypothesis 3. PSS = Perceived Stress scale; PSQI = Pittsburgh Sleep Quality Index.

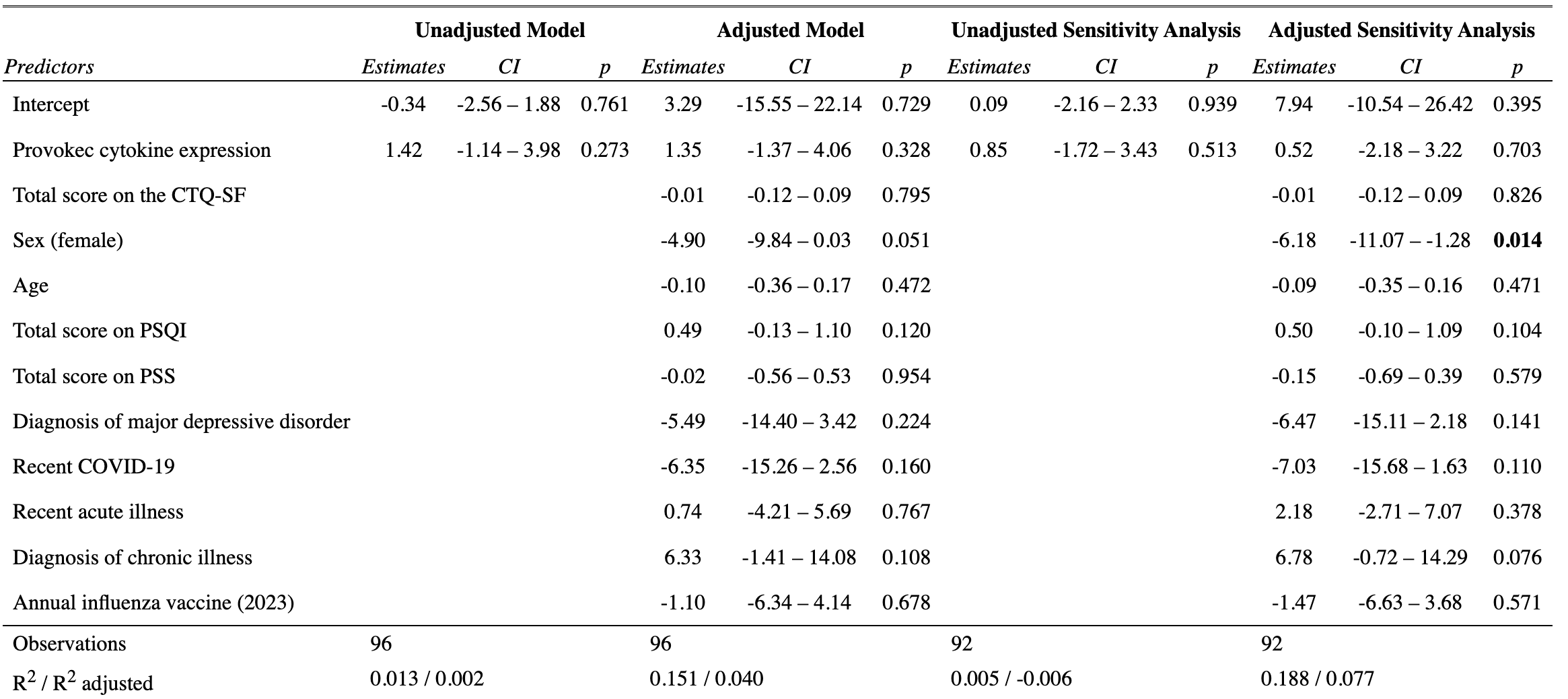

Table S18: Summary of unadjusted and covariate-adjusted models for hypothesis 3 (change in TS at the lumbar site) for the full sample and sensitivity analyses excluding 4 (of 95) participants who were unblinded to hypothesis 3. PSS = Perceived Stress scale; PSQI = Pittsburgh Sleep Quality Index.

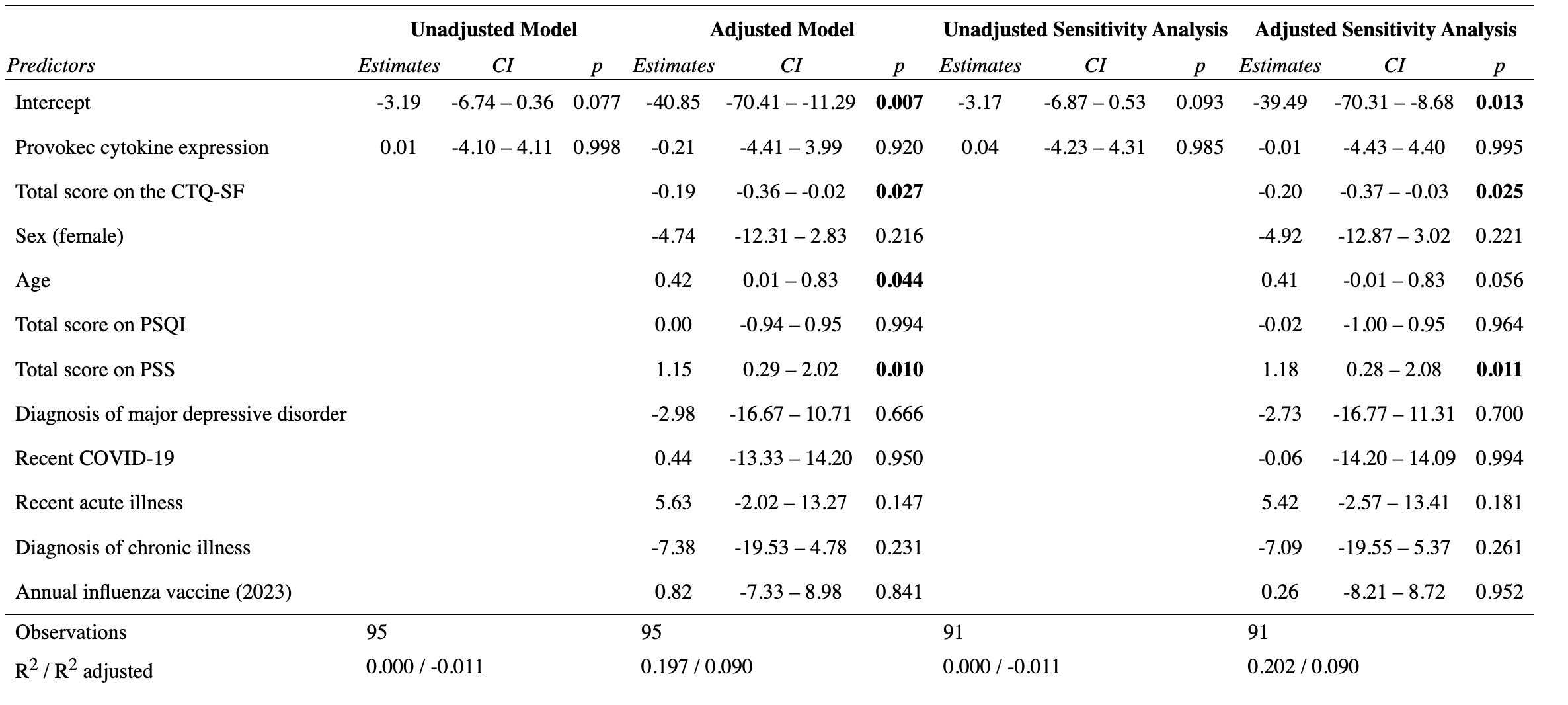

Table S19: Summary of unadjusted and covariate-adjusted models for hypothesis 3 (change in TS at the deltoid site) for the full sample and sensitivity analyses excluding 4 (of 96) participants who were unblinded to hypothesis 3. PSS = Perceived Stress scale; PSQI = Pittsburgh Sleep Quality Index.

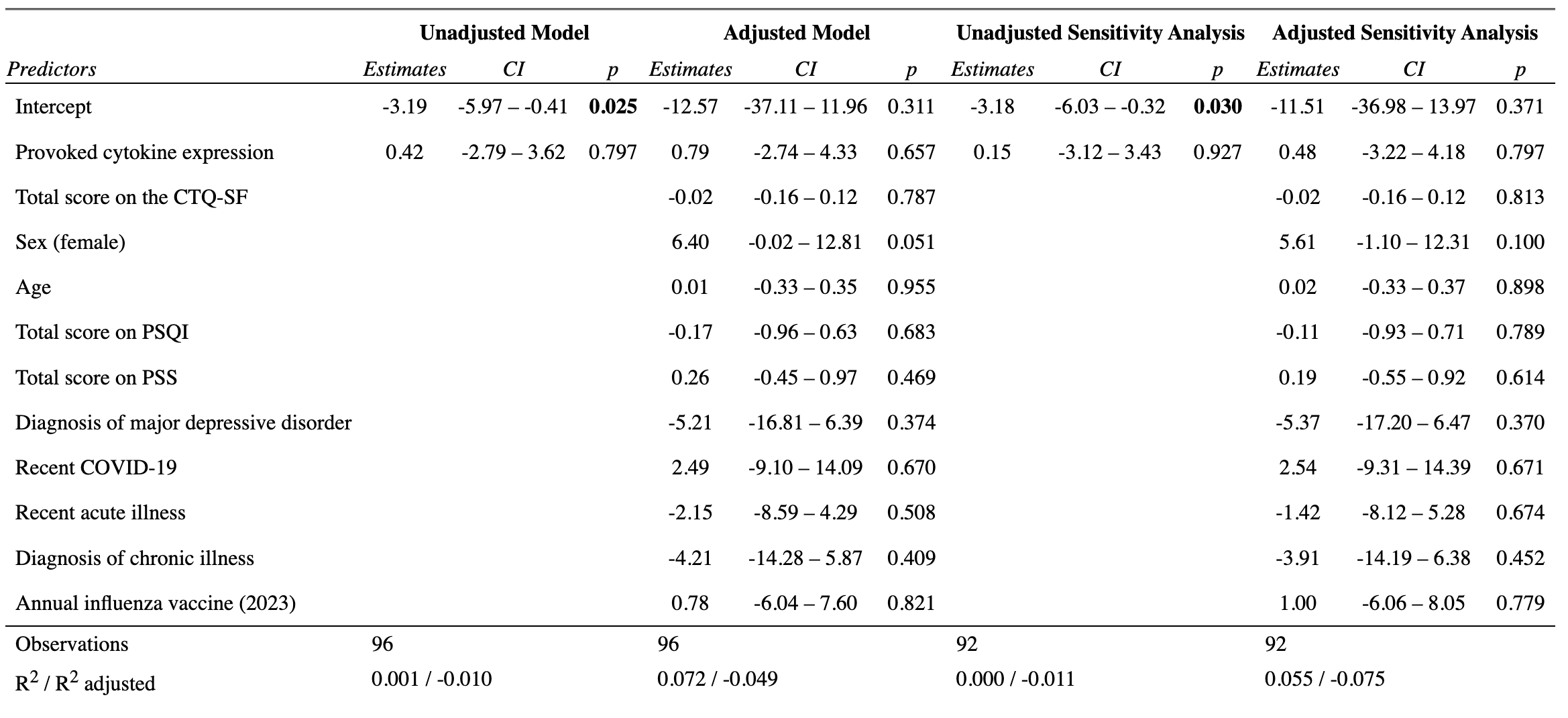

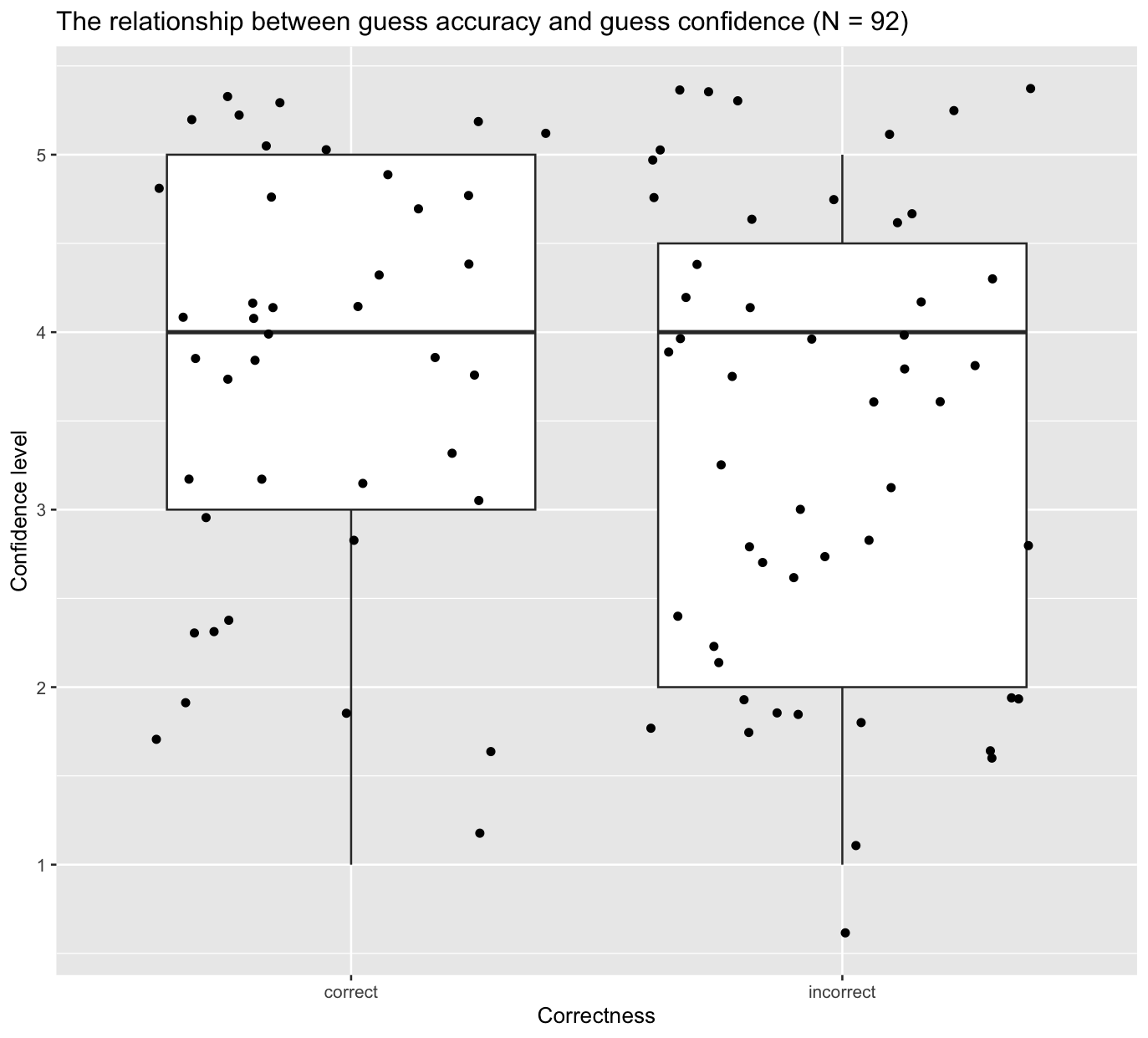

Figure S13: Blinding assessment: The relationship between the assessor’s guess accuracy and guess confidence

### 13. Exploratory analyses

#### 13.1 Relationship between provoked cytokine expression and static and dynamic light touch and single electrical stimulation

Table S20: Summary of unadjusted and adjusted models for exploratory outcomes. HFS = high-frequency electrical stimulation; PSS = Perceived Stress scale; PSQI = Pittsburgh Sleep Quality Index.

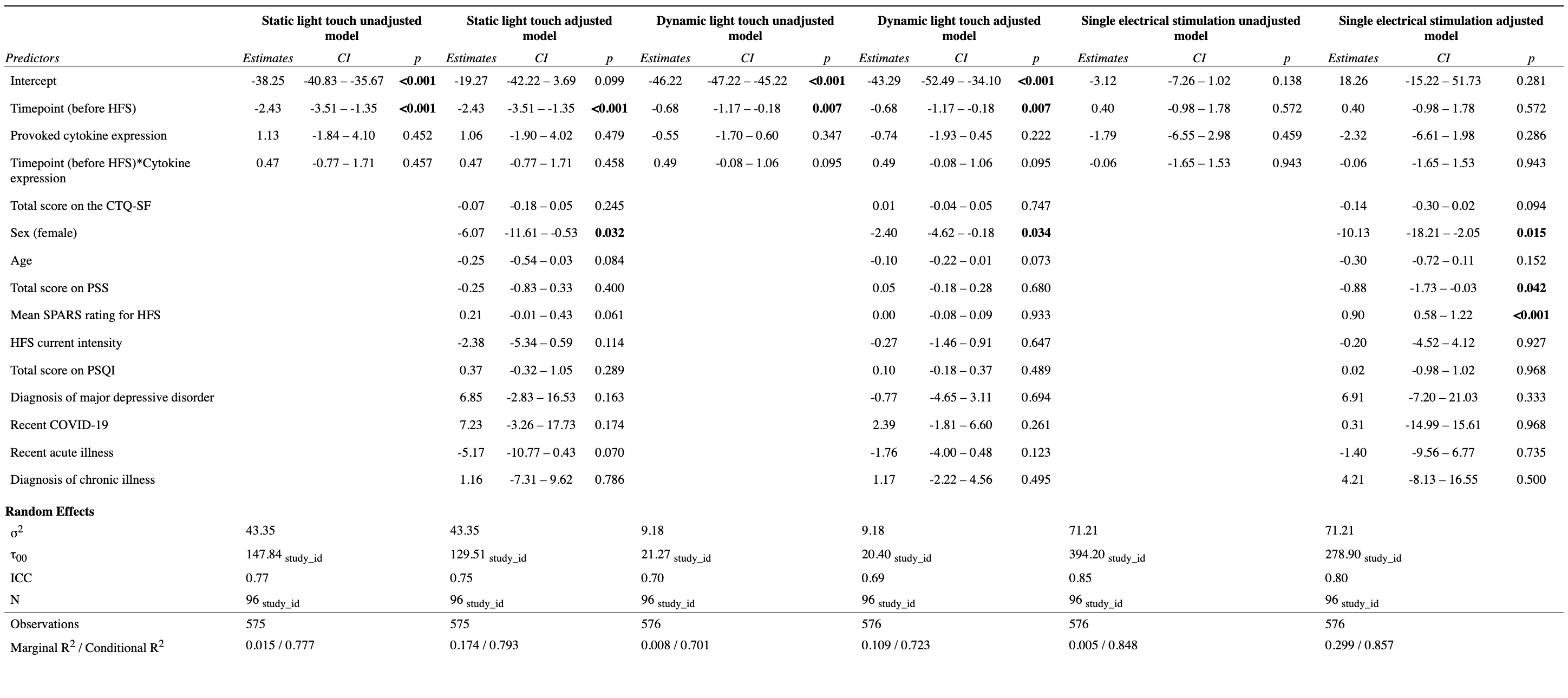

#### 13.2 Relationship between each subscale of the CTQ-SF and provoked cytokine expression

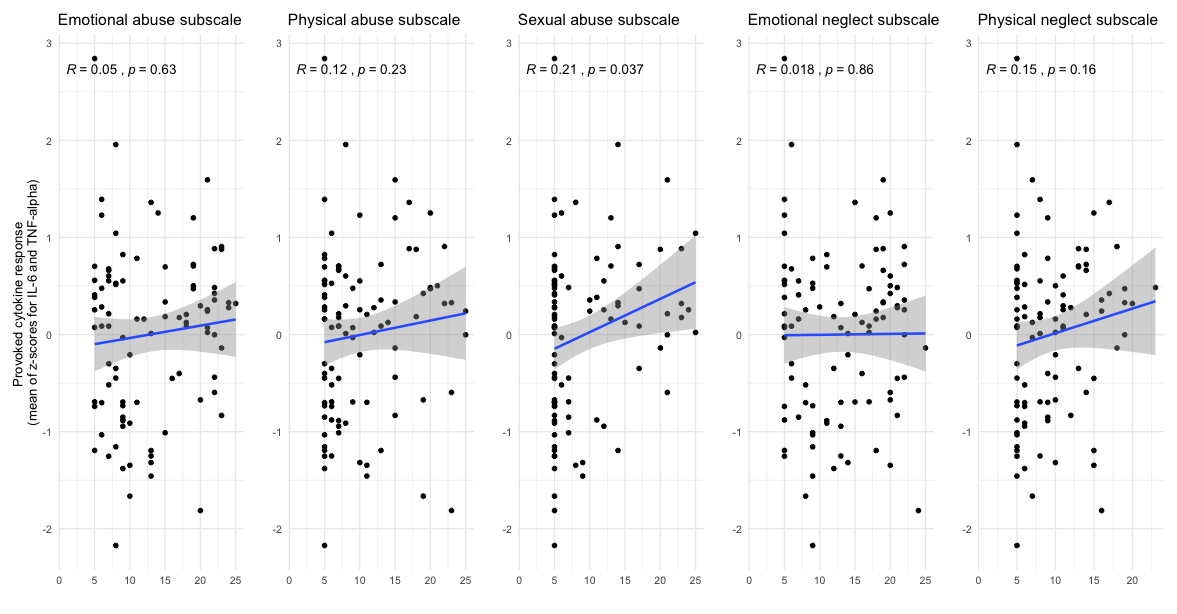

Figure S14: Correlations between each subscale of the CTQ-SF and provoked cytokine expression.

#### 13.3 Interaction between positive childhood experiences and adverse childhood experiences on provoked cytokine expression

Table S21: Summary of models for hypothesis 1 when included positive childhood experiences as a moderator in the subsample of 49 participant for whom we had data on positive childhood experiences. CTQ = Childhood Trauma Questionnaire.

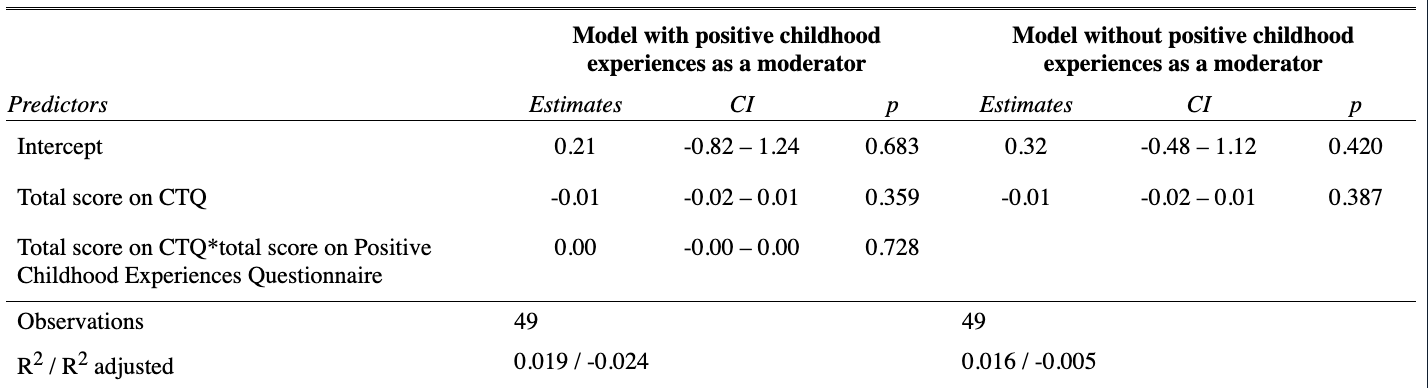
